# Individualized cortical gradient and network topology reveal symptom-linked disruptions and neurobiological subtypes in schizophrenia

**DOI:** 10.64898/2026.04.25.26351736

**Authors:** Bin Wan, Sara Larivière, Clara A. Moreau, Varun Warrier, Richard A.I. Bethlehem, Yun-Shuang Fan, Yuankai He, Ingrid Agartz, Stener Nerland, Erik G. Jönsson, Derin Cobia, Lei Wang, Benedicto Crespo Facorro, Rafael Romero-Garcia, Patricia Segura, Nerisa Banaj, Daniela Vecchio, Tamsyn Van Rheenen, Philip J. Sumner, Elysha Ringin, Susan Rossell, Sean Carruthers, Will Woods, Matthew Hughes, Gary Donohoe, Emma Corley, Ulrich Schall, Frans Henskens, Rodney Scott, Patricia Michie, Carmel Loughland, Paul Rasser, Murray Cairns, Bryan Mowry, Stanley Catts, Christos Pantelis, Aristotle Voineskos, Erin Dickie, Henk Temmingh, Freda Scheffler, Oliver Gruber, Rosanne Picotin, Vince D. Calhoun, Kyle M. Jensen, Filip Španiel, David Tomecek, Raymond Salvador, Andriana Karuk, Edith Pomarol-Clotet, Tilo Kircher, Lea Teutenberg, Frederike Stein, Udo Dannlowski, Dominik Grotegerd, Tiana Borgers, Tim Hahn, Rebekka Lencer, Carlos López-Jaramillo, Melissa Green, Yann Quide, Vaughan Carr, Stefan Ehrlich, Peter Kochunov, Christian Sorg, Melissa Thalhammer, David Glahn, Amanda Rodrigue, Kang Sim, Ali Saffet Gonul, Aslihan Uyar Demir, Nicolas Crossley, Alfonso Gonzalez-Valderrama, Philipp Homan, Wolfgang Omlor, Giacomo Cecere, Felice Iasevoli, Giuseppe Pontillo, Raquel Gur, Ruben C. Gur, Kosha Ruparel, Theodore D. Satterthwaite, Scott Sponheim, Caroline Demro, Young Chul Chung, Soyolsaikhan Odkhuu, Albert Yang, I-Jou Chi, Ole Andreassen, Lars T. Westlye, Unn K. H. Haukvik, Nadine Parker, Jakub Kopal, Dag Alnaes, Jaroslav Rokicki, Carl M Sellgren, Maria Lee, Stefan Borgwardt, Mihai Avram, Taeyoung Lee, Hang Joon Jo, Irina Lebedeva, Alexander Tomyshev, Marco De Pieri, Foivos Georgiadis, Stefan Kaiser, Paul M. Thompson, Theo G. M. van Erp, Jessica A. Turner, Boris C. Bernhardt, Sofie L. Valk, Matthias Kirschner

## Abstract

Schizophrenia is conceptualized as a brain network disorder, yet the organizational principles and heterogeneity underlying widespread cortical abnormalities remain poorly understood. Leveraging multisite MRI data from 3,958 individuals diagnosed with schizophrenia and 5,489 neurotypical individuals, we studied the cortical organization and its subtyping by analyzing individualized cortical network similarity. We used eigenvector decompositions to study spatial patterning of the gradients and graph theory to study small-world topology. Individuals with schizophrenia showed widespread alterations of gradient loadings, which followed inferior-superior and frontal-temporal axes. Alterations in small-world topology were localized in key network hubs, including the insula and anterior cingulate cortex. Multivariate brain-symptom analyses revealed a latent dimension reflecting global symptom severity, with graded contributions, strongest for disorganization and weakest for affective symptoms. This dimension was specific to gradient and topology measures, with cortical thickness showing no regional associations and an inverted symptom ordering. Finally, clustering cortical alterations identified two robust subtypes, characterized by divergent anterior cingulate (S1) versus temporoparietal (S2) thickness differences aligned with the intrinsic gradient-topology patterns. Both subtypes were present early in the illness and stable across disease stages and age groups, with S1, characterized by anterior cingulate involvement, showing higher positive symptom burden at older ages. These findings reveal systematic disruptions of cortical organization in schizophrenia, providing a network-level framework for macroscale brain organization and inter-individual heterogeneity.

## Introduction

Schizophrenia is a severe mental disorder characterized by heterogeneous symptom dimensions, including positive, negative, affective, and cognitive symptoms, and highly variable illness trajectories 1–6. Neuroimaging studies consistently demonstrate widespread cortical thickness alterations ^7–11^ with variations related to symptom profile and illness stage ^7,12–14^. This heterogeneity extends beyond clinical presentation to the brain itself: patterns of cortical alteration vary markedly across individuals with schizophrenia ^6,15–22^. Recent work has begun to capture this neuroanatomical diversity by identifying two subtypes of schizophrenia, characterized by predominant frontal versus temporal–hippocampal alterations, with morphometric differences in both subtypes subsequently extending across the cortex ^12^. Moreover, gray-matter heterogeneity appears most pronounced near illness onset and diminishes with chronicity ^23^.

Advances in network neuroscience have demonstrated that cortical changes are not isolated phenomena but are embedded within large-scale patterns of inter-regional covariance ^10,24–29^. This distinction matters conceptually: whereas conventional case-control analyses of regional cortical thickness quantify how much a given region differs between patients and controls, they do not characterize how that region’s structural variation is coordinated with the rest of the cortex ^25,29,30^. Two complementary network frameworks can be used to interrogate this coordination. First, eigenvector decomposition approaches capture continuous gradients along which cortical regions are organized according to shared covariance profiles, reflecting fundamental principles of the cortical organization ^31,32^. In healthy young adults, dominant gradients of cortical thickness covariance align with anterior–posterior and inferior–superior axes of cortical organization ^30^. Second, graph-theoretic analyses of structural covariance characterize network topology, including small-world organization, which captures the balance between local specialization and global integration ^25,33,34^. Deviations from this balance are interpreted as network disruptions and have been linked to a variety of psychiatric and neurological disorders ^29,35,36^. Because these gradient and topology measures are derived from the covariance structure across regions rather than from any single region’s magnitude of change, they can, in principle, index disruptions of macroscale cortical organization that are not reducible to regional thickness differences alone ^37^. For example, two individuals with similar regional thickness deficits could nonetheless differ in how those deficits are embedded within the broader cortical hierarchy, a distinction that gradient and topology measures, but not thickness values themselves, are designed to capture ^29,30,36,37^.

Through neurodevelopment, coordinated cortical changes follow systematic trajectories informed by evolutionary principles ^38^. For example, tangential versus radial patterns of progenitor cell division in the embryonic cortex ^39^ implies large-scale anterior-posterior and inferior-superior gradients of cortical thickness across species ^30^. Beyond early development, cortical structure continues to change across the lifespan ^40^ and is shaped by shared genetic influences across distributed regions, resulting in coordinated variation in cortical features such as thickness and area ^41,42^. Together, these findings suggest that deviations in local cortical properties are unlikely to occur in isolation, but instead reflect structured, network-level alterations constrained by developmental/genetic architecture. To elucidate how these organizational principles are perturbed in schizophrenia, it is therefore critical to characterize cortical organization by studying macroscale gradients and network topology^7,13,37,43,44^.

Building on these developmental and organizational principles, cortical covariation networks are increasingly studied in neurodevelopmental and psychiatric disorders, including schizophrenia ^28,37,44–46^. For example, a compressed anterior-posterior cortical gradient has been reported in early schizophrenia at the group level ^37^, while network topology analyses have revealed alterations in clustering within key hub regions such as the insula in adult schizophrenia ^44^. Furthermore, emerging inter-individual covariance approaches ^29,36^ show that deviations in cortical organization relate to clinical symptoms, highlighting their potential relevance for understanding disease heterogeneity ^47^. Together, these findings motivate an integrative framework that combines macroscale cortical gradients and network topology to characterize cortical covariation architecture and inter-individual variability in schizophrenia.

Here, we analyzed multisite brain MRI data from 9,447 individuals across 40 international sites from the ENIGMA Schizophrenia Working Group to comprehensively map individual cortical network organization in schizophrenia. We constructed group- and individual-level cortical thickness covariance and distance networks and derived macroscale cortical gradients together with small-world topological measures (**Fig. 1A–C**). First, we examined case vs. control differences across gradient and small-world topology to identify systematic disruptions of cortical organization (**Fig. 1D**). Second, we used multivariate association analyses to link cortical gradient and small-world topology to a shared, dimensional variation in clinical symptoms (**Fig. 1D**). Third, we applied a clustering approach based on alterations of gradients and small-world topology to identify reproducible cortical subtypes of schizophrenia (**Fig. 1E**). Finally, we characterized subtype-specific cortical thickness profiles across the adult lifespan and their alignment with macroscale cortical gradient-topology patterns (**Fig. 1E**).

**Fig. 1:**
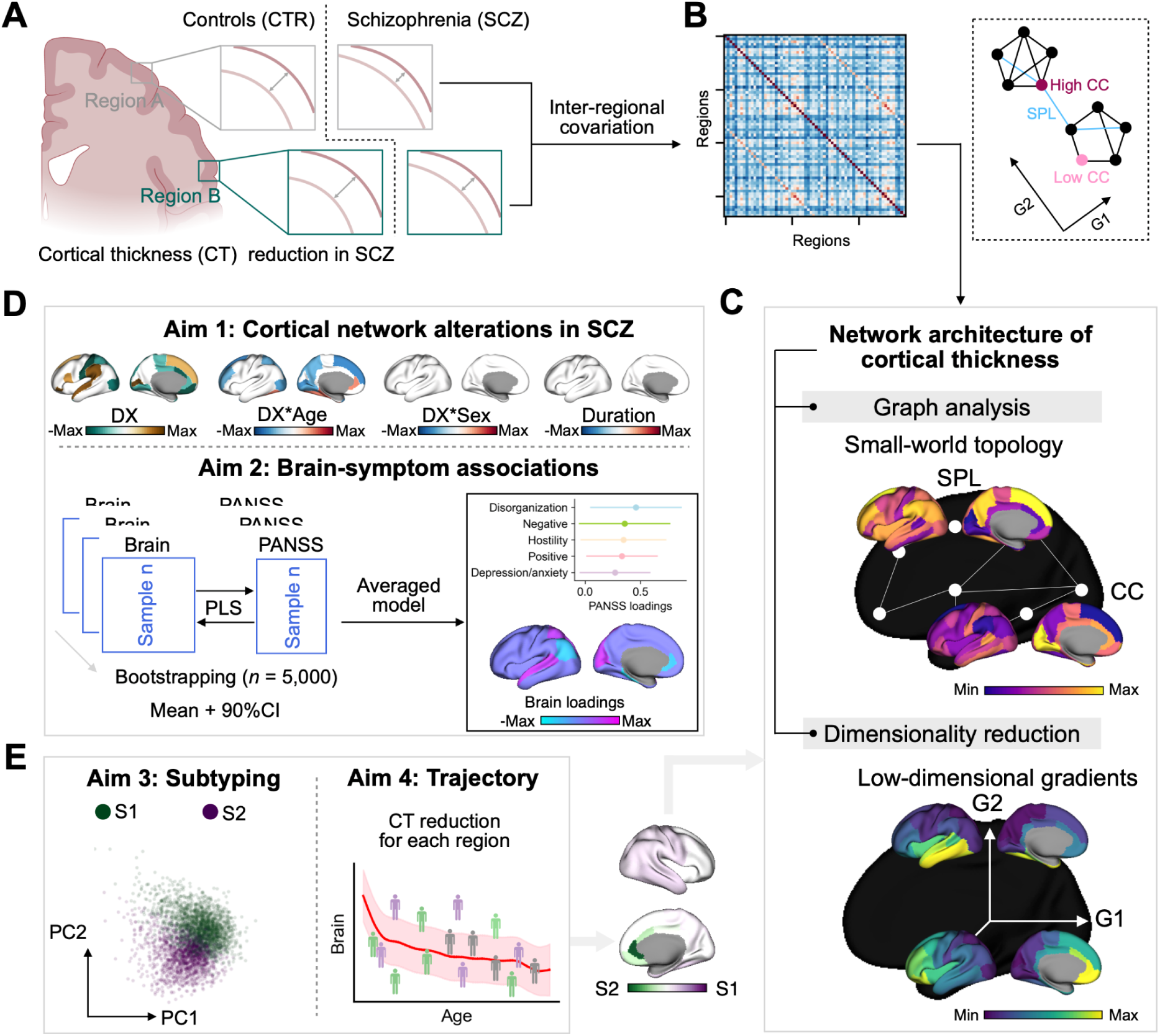
Conceptualization and aims of the study. **A:** Individuals with schizophrenia experienced extensive cortical thickness reduction. **B:** Group-level inter-regional covariance matrix is calculated by inter-subject Pearson correlation, which can be used to describe the structural network systems of cortical thickness. We can understand the matrix by reducing the dimensionality or studying the graph topology. G1 and G2 are two principal components. SPL indicates *shortest path length* between two regions and CC indicates *clustering coefficient* of a region, i.e., how many edges exist for a region. **C**: Brain maps for group-level low-dimensional gradients and small-world topology in healthy controls. All analyses were performed using the top 20% values of covariance matrix per row. **D**: In order to compare between individuals with schizophrenia and controls, we used the individual covariance formula as suggested by previous studies ^29,36^. Aim 1, focus on comprehensive characterization of cortical network alterations in schizophrenia including interaction between diagnosis (DX) and age, interaction between diagnosis (DX) and sex, and disease duration related brain maps. In aim 2, we examine the latent brain-symptom associations between cortical network architecture features and multiple clinical symptom dimensions, measured with the Positive and Negative Syndrome Scale for Schizophrenia (PANSS) using multivariate partial least square (PLS) approach with 5,000 bootstrap resamples. **E**: To address neuroanatomical heterogeneity in schizophrenia, we apply data-driven clustering to the individual-level gradient-topology profiles to uncover reproducible schizophrenia subtypes (Aim 3). We then use deviation from population modeling, to identify subtype-specific cortical-thickness profiles across the adult lifespan (Aim 4).

## Materials and Methods

### Imaging preprocessing

Following published guidelines for ENIGMA pipelines ^9,77^, all 40 participating sites processed 3D T1-weighted structural brain MRI scans using the FreeSurfer toolkit ^78,79^ and extracted cortical thickness from the standard 68 Desikan-Killiany (DK) regions ^49^. The number of scanners, vendor, strength, sequence, acquisition parameters, and FreeSurfer versions are provided in **Supplementary Materials**. Standard ENIGMA protocols for quality control were conducted at each site prior to subsequent mega-analysis (http://enigma.usc.edu/protocols/imaging-protocols).

### Structural network covariance and individual distance

Regional cortical thickness data from each individual across every ENIGMA cohort was harmonized using neuroComBat ^80^, a batch-effect correction tool that uses a Bayesian framework to improve the stability of parameter estimates. Harmonization was conducted on mean cortical thickness values with site as the primary nuisance variable (“*batch”* vector), while preserving the effects of diagnosis, age, and sex. Missing data were imputed for individuals with less than 10% missingness using IterativeImputer in the scikit-learn Python toolbox (https://scikit-learn.org/stable/index.html). Before calculating group-level structural covariance networks for schizophrenia and controls, we regressed out the effects of sex and age from the harmonized cortical thickness data. Group-level covariance matrices were then computed separately for schizophrenia and control groups. For individual-level analyses, consistent with previous studies ^29,36^, we constructed regional difference matrices by normalizing cortical thickness values relative to the control group mean and standard deviation. The formula is shown in **Fig. 2A**.

**Fig. 2:**
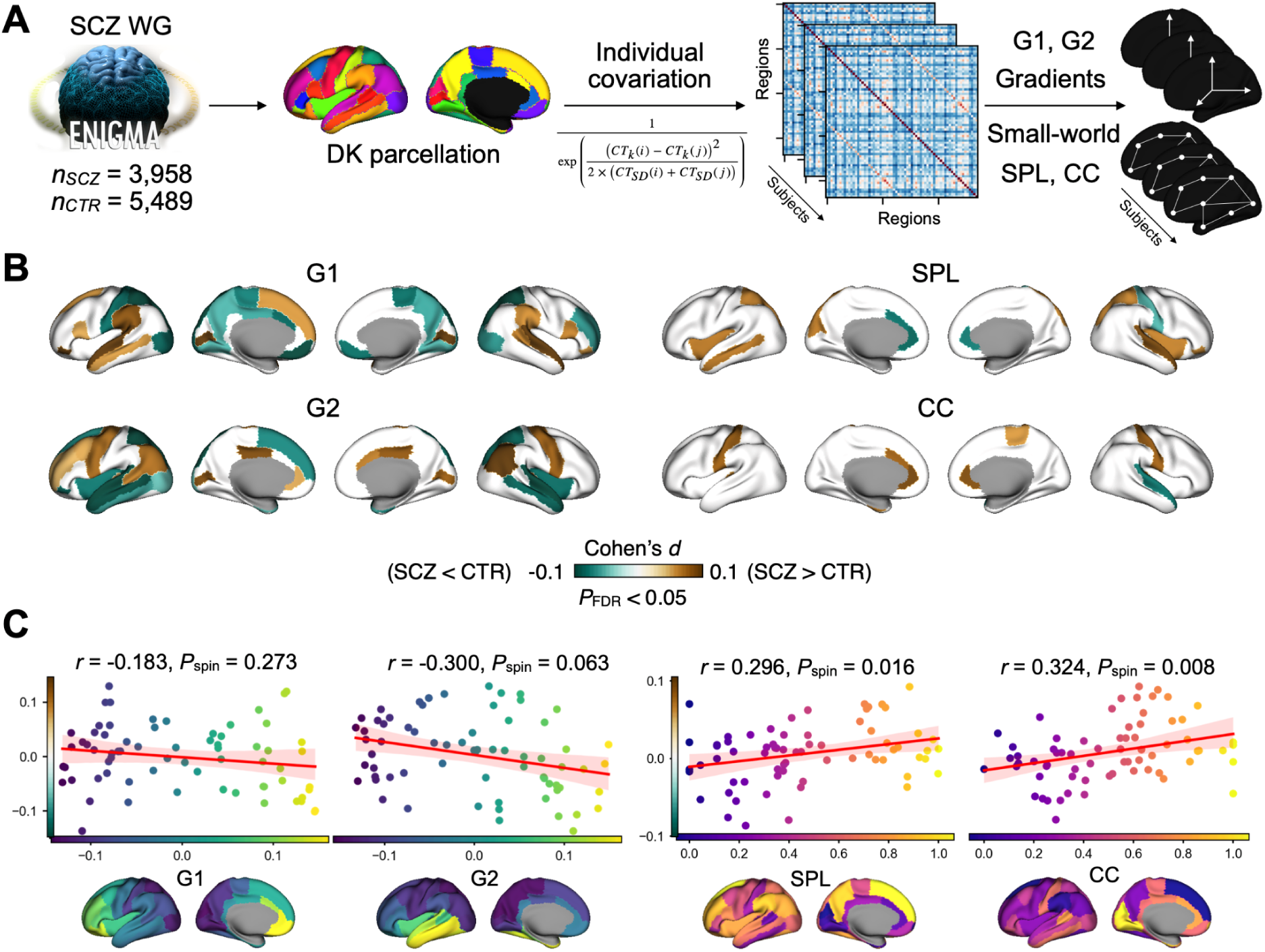
Schizophrenia-related alterations in cortical thickness network architecture. **A**: We extracted FreeSurfer outputs and demographics from the ENIGMA-Schizophrenia (SCZ) consortium including 40 data sites. Cortical thickness (CT) data were parcellated in 68 regions using Desikan-Killiany (DK) atlas ^49^. The individual variation formula was used to construct the matrix and the individual gradients (G1 and G2) and small-world topology (SPL and CC) were calculated. **B**: Cohen’s *d* brain maps were shown with the threshold of *P*_FDR_ < 0.05 from case-control comparisons. **C**: Spatial correlations between case-control Cohen’s *d* in Fig. 2B and the group-level template maps in Fig. 2C.

### Organization gradients

Given previous studies have used different sparsity densities ranging from 8% to 40% ^30,35,36,48^, we sparsified the group-level covariance matrices across thresholds from 10% to 40%, in 1% increments, to evaluate group-level template in controls as well as in SCZ. Regarding the case-control comparisons using individual difference matrices, we tested sparsity of 10%, 20% (main analysis in the Results), 30%, and 40%. Organization gradient axes were obtained using diffusion map embedding on the affinity matrix of the sparse covariance matrix using Python toolbox BrainSpace ^31,32,81^. We used alpha = 0.5 to balance the density and geometry of the matrix, which approximates the Laplace–Beltrami operator of the underlying manifold ^81^.

This approach was described by ^31^, who used a functional connectivity matrix. When applying this approach to cortical thickness ^30^, a previous study has discovered two important axes: anterior-posterior and inferior-posterior, aligning with dual origins of cortical development. In our study, we can replicate the two axes in controls and schizophrenia. We then applied a sparsity threshold of 0.8 (retaining the top 20% of covariance values) to compute individual cortical gradients, consistent with a prior ENIGMA study ^48^. To make individuals comparable, we used Procrustes rotation to align individual gradients to the group-level template generated from control data. The first two gradients (G1 and G2) were then used for subsequent analyses, which accounted for 20.4% and 17.2% of variance in controls, and 21.7% and 17.7% in schizophrenia, respectively.

### Small-world topology

Small-world topology was characterized by two nodal indices derived from the thresholded cortical thickness covariance matrix, nodal clustering coefficient (CC) and shortest path length (SPL), computed for each cortical region. CC is a regional averaged score of edges and SPL is computed for each node as the mean shortest path length to all other nodes in the network. Regional SPL is interpreted as a measure of network integration, complementary to the CC, which reflects local segregation. To make individuals comparable, scores were normalized to 0-1 for each individual. As informed by previous work, three different states were used to characterize topology, including: regular (high CC and long SPL), small-world (balance between CC and SPL), and randomized (low CCand short SPL) ^35,36^.

### Case-control comparisons

In the case-controls comparison, we used linear regression models with *brain ∼ age + sex + DX* to obtain the statistical estimates. A false discovery rate (FDR) ^82^ correction (*q* < 0.05) was then applied for brain features within each metric (G1, G2, SPL, and CC), separately. Furthermore, ICV was included as a covariate in separate models to test robustness of the group effects (**Supplementary Fig. S4)**. Interactions between diagnosis and age/sex were performed using *brain ∼ age + sex + DX + age × DX* and *brain ∼ age + sex + DX + sex × DX*. Disease duration effects in schizophrenia were examined using *brain ∼ age + sex + duration*.

### Brain-symptoms association

Brain-symptom associations were examined in the subsample of individuals with schizophrenia with available item-level PANSS and illness duration data (*n* = 624), representing 15.8% of the full schizophrenia sample (n=3,958). All data were regressed age, sex, and disease duration. Partial least squares (PLS) was used to associate clinical symptoms with brain features in schizophrenia ^83^. The significance of each latent variable was assessed by bootstrap resampling testing (*n* = 5,000), with all reported latent variables significant at *P* < 0.001. Bootstrap resampling was used to assess the reliability of individual feature loadings contributing to these significant latent variables. We report feature-level contributions at the 95% confidence interval to the established brain-symptom association. To test that results were not driven by regressing out disease duration, we repeated the PLS with regression out only regression age and sex (*n* = 881). Clinical symptoms are presented at the factor level in the main figures and at the item level in the supplementary figures. PANSS symptoms were grouped according to the five-factor model reported by ^50^. The following dimensions were identified positive (6 items), negative (8 items), cognitive/disorganization (6 items), depression/anxiety (3 items), and hostility (5 items) using exploratory factor analysis in a large multi-ethnic schizophrenia sample and validated against a meta-analysis of published PANSS models. Two PANSS items were not assigned. Finally, given that the main analysis used a single component, we additionally fitted a three-component solution to test for further structure. Because PLS components are extracted in order of decreasing explained covariance, the absence of significant loadings on the second and third components indicates that higher-order components would not be informative, and the analysis was therefore not extended further.

### Subtyping

A *k*-means algorithm was used to cluster SCZ individuals using significant brain features and the SSE was calculated for each different cluster solution (2 through 9). However, no clear convergence was observed in the SSE curve, therefore, principal components analysis (PCA) was applied to reduce the dimensionality of the brain features;the first two PCs explained 9.18% and 6.13% of the total variance, respectively (**Fig. 3B**). In the space defined by the first two PCs, a two-subtype solution provided the best separation. The sample was then split into two subsamples for cross-validation of the subtyping approach, yielding a high assignment concordance in both subsamples (> 98%). We subtyped the subjects using both significant brain features (main) and all features (supplement).

**Fig. 3:**
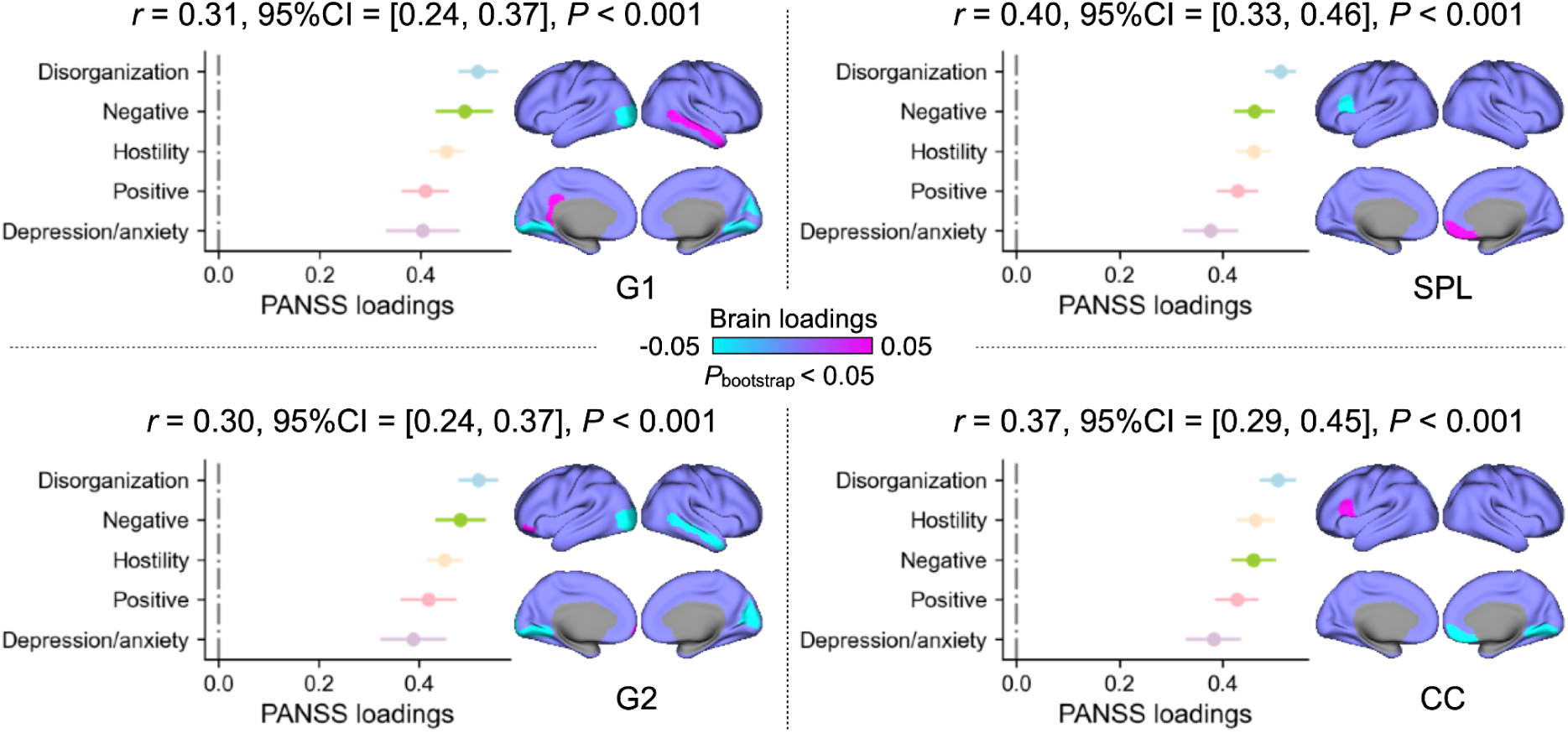
Dimensional associations with clinical profiles. Multivariate brain-symptom associations (PLS: partial least squares regression) for G1, G2, SPL, and CC separately were conducted. We selected the averaged model over 5,000 bootstrap resamples. We used 95% confidence interval (CI) to test the significance of brain and clinical features, which is *P* < 0.05.

### Normative models

Generative additive models (GAM) were used to generate the population models and deviation scores using the Python toolbox pyGAM (https://pygam.readthedocs.io/en/latest/). Models were fit in controls, with age specified as a smooth term, sex as a categorical factor, and ICV as a linear covariate, to estimate mean and regional cortical thickness. For each region, we derived the predicted mean and 95% CI, and computed deviation scores as *(observed - predicted cortical thickness) x 4 / (upper - lower 95% CI)*, approximating a *z*-score.

### Spatial autocorrelation null models

To account for spatial autocorrelation between cortical maps, we used spin permutation tests to obtain corrected *P*-values ^32^. The DK cortical values were projected onto the fsaverage5 surface and the surface was randomly rotated 1,000 times to generate surrogate maps. Null distributions were constructed from these rotations, and *P*-values were calculated as the proportion of null models exceeding the empirical statistic. Analyses were conducted using the ENIGMA toolbox ^84^.

## Results

### Data sources

We studied 3,958 individuals with schizophrenia (SCZ; 1,488 females, age mean ± SD = 35.0 ± 12.2 years) and 5,489 healthy controls (CTR; 2,783 females, age mean ± SD = 34.6 ± 13.40 years) from 40 centers of the international ENIGMA consortium Schizophrenia Working Group. Diagnosis of schizophrenia or schizoaffective disorder was confirmed at each center using criteria from either the International Classification of Diseases (ICD) or Diagnostic and Statistical Manual (DSM). Details on site-specific demographic and imaging information are provided in **Supplementary Materials**.

### Schizophrenia-related alterations in cortical thickness network architecture

We characterized cortical network architecture using principal gradients of dimensionality reduction and small-world topology derived from cortical thickness similarity matrices. We first performed group-level analysis of the covariance matrix to obtain the first two gradients (G1 and G2) and small-world topology (SPL and CC) using sparsity scores ranging from 0.6 (top 40% remained) to 0.9 (top 10% remained) with a 0.01 interval (**Supplementary Fig. S1**). Two principal gradients describing cortical thickness covariance along anterior-posterior (G1) and posterior-temporal (G2) axes were identified, with their order varying by sparsity level and group **(Supplementary Fig. S2**). The temporal anchor of G2 extended into occipital regions, largely overlapping with the anterior–posterior axis described in normative structural covariance studies (Valk et al., 2020). SPL and CC showed reversed spatial patterns, with higher normalized SPL in frontal regions and higher normalized CC in visual regions (**Supplementary Fig. S3**). For all subsequent analysis, we chose a sparsity of 0.8 as it minimized similarity between G1 and G2 (**Supplementary Fig. S1**), and aligns with thresholds used in previous studies ^35,36,48^. Individual-level gradients were computed using subject-specific difference matrices and aligned to control group-level templates via Procrustes rotation, while individual topology metrics were rescaled to a 0-1 range (**Fig. 2A**).

Individuals with schizophrenia exhibited system-wide alterations along cortical network gradients and more localized disruptions in small-world topology (**Fig. 2B**, all *P*_FDR_ < 0.05). For G1, alterations were observed in superior temporal, postcentral, occipital, and prefrontal regions. The observed gradient alterations did not spatially align with the G1 axis and showed a widespread, though randomly distributed, pattern (Pearson *r* = -0.183, *P*_spin_ = 0.273, **Fig. 2C**). In contrast, alterations along the second gradient (G2), reflecting the frontal-temporal cortical axis, followed a systematic spatial pattern. Affected regions included the left lateral occipital gyrus, bilateral lingual gyrus, superior temporal sulcus and gyrus, precentral gyrus, insula, medial prefrontal areas, and middle cingulate cortex; this disruption pattern closely mirrored the normative structural covariance organization of G2 but without statistical significance (Pearson *r* = -0.300, *P*_spin_ = 0.063, **Fig. 2C**). Thus, while G1 alterations were randomly distributed along the anterior-posterior axis, G2 alterations showed a more structured disruption pattern along the posterior–temporal axis. When compared with the case-control cortical thickness map, gradient alterations showed no significant spatial correlation (G1: Pearson *r* = -0.201, *P*_spin_ = 0.171; G2: Pearson *r* = 0.198, *P*_spin_ = 0.121). Detailed regional statistics are reported in **Supplementary Materials**. Small-world topology disruptions were more spatially constrained but located in key integration hubs. SPL alterations concentrated in the bilateral anterior cingulate cortex, left insula, and superior parietal cortex, while CC changes affected anterior cingulate and postcentral regions. Both topological measures showed significant spatial correlations with their respective normative network patterns (SPL: Pearson *r* = 0.296, *P*_spin_ = 0.016; CC: Pearson *r* = 0.324, *P*_spin_ = 0.008), indicating systematic disruption of local network efficiency (**Fig. 2C**). In contrast to the gradients, topological disruptions showed moderate spatial correspondence with the case–control cortical thickness map (SPL: Pearson *r* = -0.455, *P*_spin_ = 0.002; CC: Pearson *r* = 0.432, *P*_spin_ < 0.001). Detailed regional statistics are reported in **Supplementary Materials**.

We assessed the robustness and sensitivity of cortical network alterations across analytical parameters, including covariate adjustments, sparsity thresholds, and demographic factors (age, sex and duration of illness). First, gradient and topology alterations remained highly consistent with spatial correlation maps between non-ICV and ICV-controlled Cohen’s *d* maps exceeding *r* > 0.98 (**Supplementary Fig. S4**). We further replicated the diagnostic effects of different sparsity parameters (10%, 30%, and 40% of covariance edges, **Supplementary Fig. S5**), where the overall spatial patterns of Cohen’s *d* were similar. Gradients-related regions remained consistent across thresholds, although additional regions emerged at lower sparsity level (i.e., denser matrices). We also examined age × diagnosis (DX) and sex × DX interactions, as well as effects of disease duration **Supplementary Fig. S6**. Significant age × DX interaction along G1 were observed in bilateral occipital and superior parietal cortices, as well as the left prefrontal and anterior cingulate cortex. For G2, age × DX interactions were observed in bilateral prefrontal, superior parietal, and occipital cortex; no regions survived FDR correction for SPL and CC. For sex × DX interactions, no regions survived FDR correction in G2, SPL, or CC, only one region (left precentral gyrus) was significant for G1. No regions showed significant effects of disease duration.

### Dimensional associations with symptom profiles

To determine how cortical network architecture relates to clinical heterogeneity in schizophrenia, we examined associations with symptom dimensions using multivariate partial least squares (PLS with one component) analysis in the subsample with available item-level PANSS data (*n* = 624, **Fig. 3**). In the main analysis, age, sex, and disease duration were regressed out from all brain and symptom features. Disease duration was specifically included as a covariate because symptom profiles vary systematically with illness duration in schizophrenia ^1–6^, and removing this variance isolates the brain–symptom associations of interest. Both gradients and topology showed robust significant associations with global clinical symptom burden across all five PANSS factors: positive, negative, cognitive/disorganization, depression/anxiety, and hostility ^50^. The overall brain-symptom association was robust at the model level (*P*_bootstrap_ < 0.001), with testing the 95% bootstrapping confidence interval (CI) as significance. The strength of these associations followed a clear hierarchy, with depression/anxiety showing the weakest loadings and disorganization the strongest. Regionally, G1 and G2 converged on the right middle temporal cortex and occipital cortex, but each also showed distinct positive loading patterns: G1 was additionally associated with the left posterior cingulate cortex, whereas G2 was additionally associated with the inferior frontal cortex (**Fig. 3)**. Regarding topology, SPL and CC showed significant loadings on the left inferior frontal and medial prefrontal cortices. These findings suggest that both gradients and disrupted network efficiency are directly related to the clinical manifestations of schizophrenia. The robustness of the network-symptom relationships was further supported by item-level analysis, which preserved both the clinical loading hierarchy and regional specificity (**Supplementary Fig. S7**). Finally, repeating the analyses with regressing out age and sex only revealed comparable findings (**Supplementary Fig. S8**).

To test whether these brain-symptom associations were specific to cortical network organization rather than reflecting underlying regional cortical thickness differences,, we repeated the PLS analysis using cortical thickness. In the model regressing age and sex only, no significant symptom or brain loadings were identified. In the model additionally regressing disease duration, no significant brain loadings emerged, yet all symptom dimensions except disorganization reached significance. Critically, the rank order of symptom loadings was inverted relative to the gradients/topology model (**Supplementary Fig. S9**). Together, these results indicate that gradient and topology measures are associated with a distinct symptom dimension compared to cortical thickness, and were more sensitive in detecting regional brain-symptom associations.

Because PLS with one component restricts all symptoms to a single axis, we tested whether additional components would reveal further separable brain-symptom patterns by repeating the PLS with three components. PLS1 yielded a similar pattern to the one-component solution, while PLS2 and PLS3 showed no significant brain or symptom loadings (**Supplementary Fig. S10**), indicating that there is no reliable structure beyond the first component. This supports a single general symptom dimension linked to regional gradient and topology measures rather than multiple dissociable brain-symptom axes.

### Two subtypes identified by altered network architecture in schizophrenia

Next, to study the heterogeneity of the systematic disruptions of cortical organization in schizophrenia, we clustered these individuals using 82 significant features identified in the case–control analysis from **Fig. 2B**, which delineates biologically meaningful subtypes in schizophrenia (*n* = 3,958, **Fig. 4A)**.

**Fig. 4:**
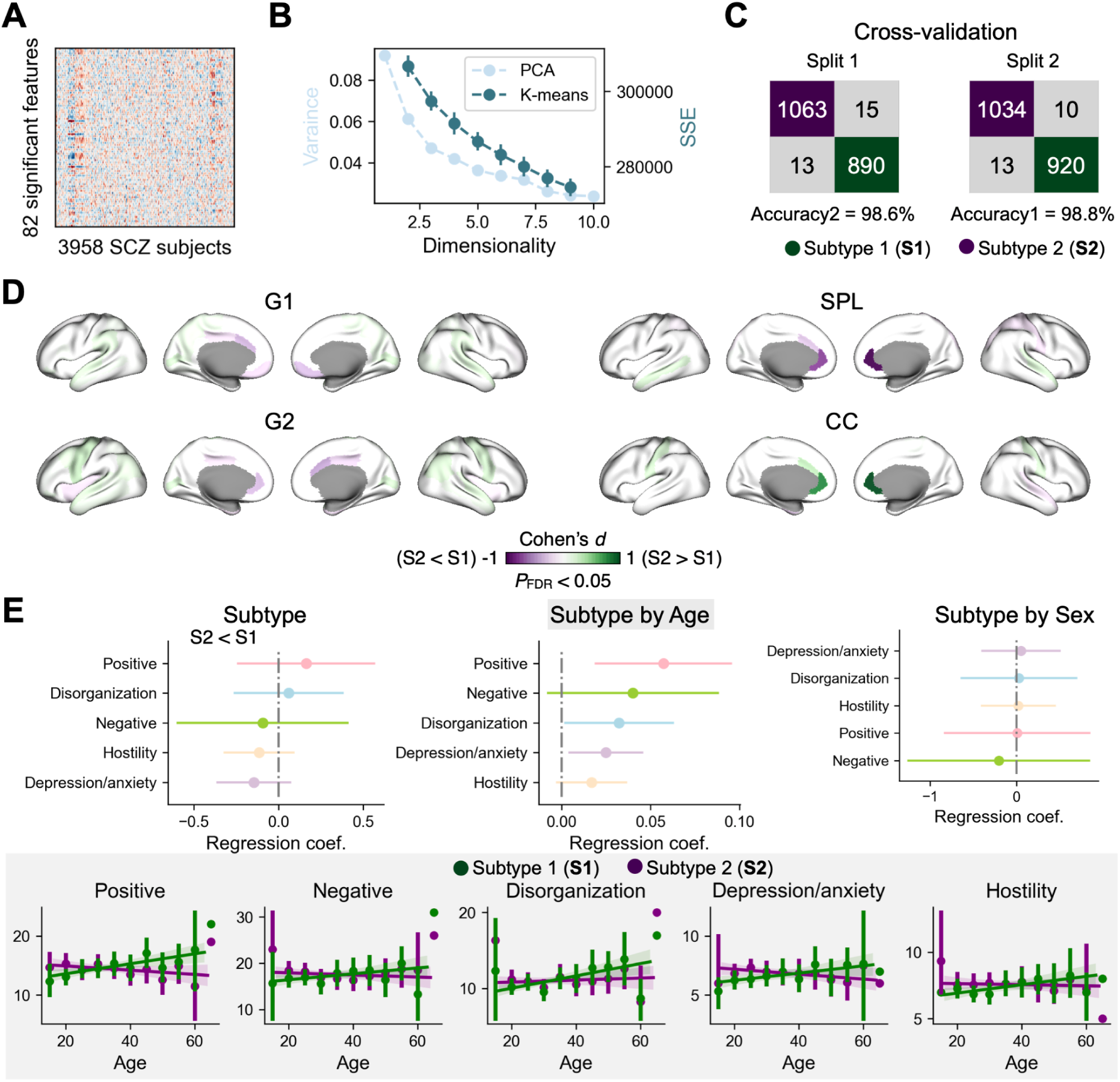
Subtypes in schizophrenia. **A**: The input matrix of 82 significant features from case-control comparisons in Fig. 2B in 3,958 individuals with schizophrenia. **B**: Principal component analysis (PCA) was performed on the matrix to reduce the dimensionality of brain features. The first two PCs explained 9.9% and 9.2% of the total variance. K-means was performed on the matrix to cluster subjects into subtypes. K cluster number ranges from 2 to 9. Sum of squared errors (SSE) indicate the sum of distance between each data point and its assigned centroid. **C**: Cross-validation of the two subtypes (S1 and S2) by 1:1 splitting the whole sample into two subsamples. Both assignment concordances were greater than 98%. **D**: Subtype difference in gradients and small-world topology (82 features from **A**). **E**: Subtype difference in schizophrenia clinical factors as well as interactions of subtype with age and sex. We additionally showed the age correlation with symptom profiles in S1 and S2 separately given the significant subtype-by-age interactions.

A *k*-means clustering algorithm was first applied to this individual-by-feature matrix (**Fig. 4B**), but the sum of squared errors (SSE) curve did not show clear convergence along 2–9 clusters. To improve separability, we reduced feature dimensionality using principal component analysis (PCA, **Supplementary Fig. S11**) to help identify the ideal number of subtypes. Overall, principal components 1-5 accounted for 9.18%, 6.13%, 4.70%, 4.20%, and 3.63% of the total variance. In this space, a two-subtype solution (S1 and S2) showed a clear separation of the data from the scatter plot (**Supplementary Fig. S11**). Robustness of the two subtypes was confirmed by half-split cross-validations, yielding assignment concordances of 98.6% and 98.8% across splits **(Fig. 4C**). In total, 2,125 individuals (53.7%) were categorized as S1 and 1,833 (46.3%) as S2. There were no significant differences in age (*t* = 1.68, *P* = 0.093), sex ratio (chi-square = 0.66, *P* = 0.415), or disease duration (*t* = 0.62, *P* = 0.532) between subtypes.

We observed significant subtype differences across all 82 brain network features (**Fig. 4D**) mirroring the case-control difference in gradient and network topology observed when comparing the entire schizophrenia sample with controls (**Fig. 2B**). Direct comparison between S1 and S2 revealed that S2 exhibited a more schizophrenia-like pattern while the pattern of S1 was relatively similar to the control group (see **Fig. 2B)**. Subtypes did not differ in the five symptom factors. However, in the linear regression of *symptom ∼ age + sex + subtype + age × subtype*, there were significant interactions between subtype and age in positive (*t* = 2.95, *P* = 0.003), disorganization (*t* = 2.10, *P* = 0.036), and depression/anxiety symptoms (*t* = 2.40, *P* = 0.016), shown in **Fig. 4E**. In particular, S1 showed a positive correlation between age and symptoms (positive: Pearson *r* = 0.146, *P* = 0.003; disorganization: Pearson *r* = 0.177, *P* < 0.001; depression/anxiety: Pearson *r* = 0.102, *P* = 0.037), whereas correlations for S2 were negative but did not reach statistical significance.

Item-level analyses revealed that subtypes did not differ on PANSS items (**Supplementary Fig. S12**). Subtype × age interactions were significant in delusions, unusual thought content, anxiety, preoccupation, conceptual disorganization, guilt, active social avoidance, lack of judgement insight, and grandiosity. No significant subtype × sex interactions were observed for any PANSS item.

Together, *k*-means clustering based on cortical network architecture features identified two subtypes that differ robustly in gradient organization and network topology, as well as age correlations with symptom dimensions.

### Cortical thickness reduction differs between two subtypes

Next, we used generalized additive models (GAM) to examine how the cortical thickness profiles of the two subtypes deviate from normative trajectories of cortical thickness across the adult lifespan. We first fitted GAM for mean cortical thickness in controls and extracted individual deviation scores (*z*-scores). Heterogeneity was defined as the proportion of individuals showing extreme deviations (i.e., |z-score| > 2). Compared to controls (*z*-score = 0.00±1.02, heterogeneity = 4.5%), both subtypes showed lower mean cortical thickness z-scores (S1: *z*-score = -0.53±1.05, heterogeneity = 9.3%; S2: *z*-score = -0.66±1.13, heterogeneity = 11.7%). Mean z-scores differed significantly between S1 and S2 (S1 vs S2: *t*= 3.63, *P* = 0.0003) as well as between each subtype and controls (S1 vs controls: *t* = 20.53, *P* = 3.49e-91; S2 vs CTR: *t* = 23.61, *P* = 7.31e-119, upper right corner of **Fig. 5A**).

**Fig. 5:**
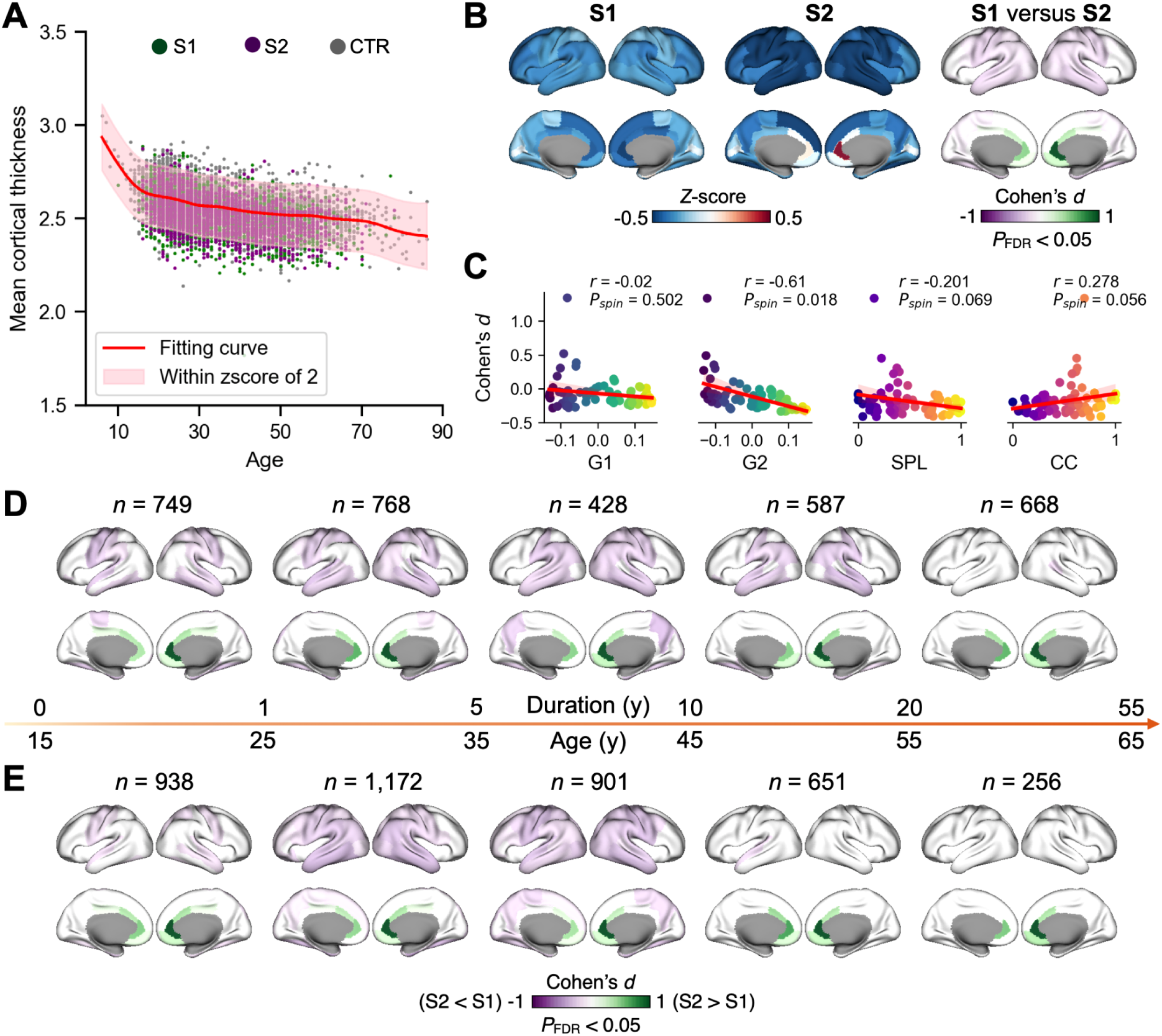
Cortical thickness profiles of subtypes. **A**: Normative model of mean cortical thickness using healthy controls data (*n* = 5,285). Deviation scores (z-scores) were derived from the normative model for each individual including schizophrenia and healthy control. **B**: Using regional normative models, we extracted z-scores for each region and individual. We compared z-scores between subtype 1 (S1) and subtype 2 (S2). **C:** We also correlated the S1 versus S2 z-score difference (Cohen’s *d*) map with the group level gradient-topology patterns from the controls in our cohort. We visualized the difference maps in different disease duration (**D**) and age (**F**) groups. Disease duration was categorized into five groups: 0-1, 1-5, 5-10, 10-20, and 20-55 years. Age was categorized into five groups from 15 to 65 years with a 10-year interval.

In a second step, we applied GAM for each cortical region to identify subtype-specific profiles of regional deviations from the normative cortical thickness patterns. In particular, S1 showed a cortical thinning pattern across the whole cortex, while S2 showed cortical thinning in medium frontal, temporal, parietal, and central cortices but increased thickness in anterior cingulate cortex (**Fig. 5B**). Comparing regional z-scores between S1 and S2, revealed that S2 showed significantly thinning in the temporal, central, and parietal regions relative to S1, while S1 showed thinning in the anterior cingulate cortex relative to S2 (**Fig. 5B**, *P*_FDR_ < 0.05). These subtype differences in regional cortical thickness were highly spatially correlated with group level gradient-topology patterns of G2 (G1, Spearman *r* = -0.020, *P*_spin_ = 0.502; G2, Spearman *r* = -0.610, *P*_spin_ = 0.018; SPL, Spearman *r* = -0.201, *P*_spin_ = 0.069; CC, Spearman *r* = 0.278, *P*_spin_ = 0.056; **Fig. 5C**).

To avoid confounding of mean thickness in the differences between subtypes (mentioned above), we performed regression analysis with controlling for mean thickness. The spatial Cohen’s *d* maps from with and without controlling for mean thickness were highly similar (Pearson *r* = 0.997, *P*_spin_ < 0.001, **Supplementary Fig. S13**). It suggests the S1 versus S2 pattern is not driven by the mean thickness.

To examine whether the cortical thickness differences between subtypes were driven by the features used for subtyping, we repeated the clustering using all network features rather than only the significant ones. The thickness differences between subtypes remained highly consistent with the original result (Pearson *r* = 0.589, *P*_spin_ < 0.001, **Supplementary Fig. S13**), indicating that the pattern was not an artefact of feature selection.

Finally, we examined whether cortical thickness differences between S1 and S2 were evident at early illness stages and whether these subtype-specific profiles relate to disease duration or age. Stratifying patients by duration of illness (0–1, 1–5, 5–10, 10–20, and 20–55 years) showed that regional z-score differences between S1 and S2 were already detectable in the earliest stage (0–1 year) and remained robust across first four subsequent duration bins (FDR-corrected; **Fig. 5C**, see **Supplementary Fig. S14C** for subtype-specific mean z-score maps for each duration bin). The observed effects in temporal and parietal regions were not significant in the last bin, but the anterior cingulate cortex finding remained. The spatial pattern of subtype differences was qualitatively consistent across stages, indicating that these cortical profiles emerge early in the illness course and persist throughout its progression. Complementary regression models testing duration by subtype interactions in mean and regional cortical thickness were not significant (*P*_FDR_ > 0.1), suggesting comparable trajectories across subtypes (**Supplementary Fig. S14A-B)**.

A complementary age-stratified analysis (15-65 years, 10-year intervals) revealed a similar pattern: regional z-score differences between S1 and S2 remained significant across the first three age bins (FDR-corrected; **Fig. 5D**, see **Supplementary Fig. S15C).** Age bins after 45 years were mainly significant in the anterior cingulate cortex. The spatial distribution of subtype differences closely resembled that observed across bins of illness-duration, and no significant interactions between age x subtype or sex x subtype were detected (**Supplementary Fig. S15A-C**). Together, these findings indicate that distinct cortical thickness profiles of S1 and S2 are established early and remain stable across illness duration, adult age range, and sex.

## Discussion

Leveraging the largest multisite individual-level dataset of schizophrenia to date (>9,000 participants from the ENIGMA Schizophrenia Working Group), we examined individual macroscale cortical network organization by integrating low-dimensional gradients with small-world network topology. Across patients, we observed system-wide alterations of the inferior-superior (G1) and the frontal-temporal (G2) axes of network organization. Cortical gradient alterations in G1 showed a largely random and spatially heterogeneous disruption, whereas alterations in G2 showed a somewhat more systematic compression along the occipital and temporal anchor. In contrast to these widespread gradient alterations, small-world topology showed more localized cases-control differences in key integration hubs including the insula and anterior cingulate cortex. Both gradients and small-world topology showed selective and robust brain-symptom associations reflecting global symptom severity, most strongly for disorganization, whereas cortical thickness showed no regional associations and an inverted symptom ordering. These findings point to (i) a core, system-wide gradient reorganization characteristic of schizophrenia, and (ii) a network component rather than local thickness accounting for individual differences in symptom expression. Finally, the data-driven clustering of the disorder-specific gradient-topology features identified two reproducible biological subtypes reflecting the whole cortex (**S1**) and (temporal, central, medial frontal, and parietal) (**S2**) patterns of reduced cortical thickness. Comparison of cortical profiles further differentiated the subtypes, with anterior cingulate predominance in S1 and posterior predominance in S2. These differential subtype-specific cortical profiles aligned along the group-level gradient-topology patterns, emerged early in the illness course (<1 year of duration), and remained detectable across the full adult age range, independent of sex. Together, these findings delineate systematic cortical network disruptions in schizophrenia, encompassing widespread gradient alterations and topology–symptom associations, alongside organization-derived subtypes with distinct cortical profiles that arise early and remain stable across the adult lifespan.

At the level of cortical organization, schizophrenia was characterized by widespread disruption of structural covariance gradients. The inferior-superior gradient (G1) displayed a largely random pattern of change, whereas the frontal-temporal gradient (G2) showed a compression-type shift in gradient organization. Developmentally, this compression may reflect delayed or incomplete maturation of anterior association cortices ^37^, consistent with neurodevelopmental models of schizophrenia emphasizing disrupted cortical organization formation and impaired prefrontal specialization ^51,52^. Functionally, diminished separation along this axis may underlie the blending of perceptual and higher-order cognitive processes characteristic of the disorder ^53^. The widespread yet spatially heterogeneous disruption along the inferior–superior axis may impair integration between ventral archicortical and dorsal paleocortical developmental streams ^30,54,55^. Within this dual-origin framework ^30^, such diffuse alterations suggest a failure to maintain the normal differentiation between these cortical lineages, consistent with models of asynchronous maturation and reduced cortical specialization in schizophrenia. From a genetic perspective, these dual cortical origins, as indexed by morphometric inverse divergence, exhibit strong genetic correlation and are linked to schizophrenia risk ^56^. This suggests that the observed gradients may be driven, at least largely, by shared genetic architecture.

In contrast to these system-wide gradient alterations, the analysis of small-world topology revealed regionally specific network disruptions centered on key cortical hubs, including the superior parietal, insula, and anterior cingulate cortices. Among these regions, insula is of particular interest, as previous studies have also identified reproducible lower network efficiency (i.e., higher SPL or lower CC) in this region in individuals with schizophrenia ^44,57^. Serving as a crucial interface between the temporal and prefrontal regions, both in anatomical terms and in its functional capabilities, insula plays a pivotal role in the discrimination between self-generated and external information ^44,58^. Notably, the broader pattern of topological disruption seen in our study including regions such as the insula, anterior cingulate and superior parietal cortices mirrors prior network-based studies that consistently implicate these regions as central nodes in schizophrenia-related cortical organization ^7,43,59^. Unlike gradient alterations, topological disruptions showed a moderate spatial correspondence with cortical thickness effects. This likely reflects the sensitivity of small-world properties to the strength of covariance edges incident on each region: regions with greater thickness reduction also show degraded local efficiency and longer path lengths, because their covariance edges are disproportionately weakened. Together, these findings extend prior work by linking localized disruptions at cortical hubs to system-level cortical organization, and by showing that these disruptions are only partially captured by regional thickness while carrying distinct clinical information.

Linking cortical gradients and topology to clinical symptoms revealed a consistent dimensional pattern in which both cortical network features were significantly associated with psychopathology. The shared symptom dimension was led by disorganization, followed by hostility, positive and negative symptoms with weaker contributions from affective domains. This distinction is clinically relevant, as negative and disorganized symptoms are typically more persistent and prognostically informative than positive symptoms ^4,60–62^, and are associated with poorer functional outcomes and prognosis ^4,61,63–65^. Rather than mapping onto isolated symptom-specific loci, the identified associations suggest a shared symptom dimension supported by gradients and topological disruption across a set of key cortical regions, including middle temporal, inferior frontal, and orbitofrontal areas. These regions are implicated across multiple studies of disorganization, positive and negative symptoms and are central to executive control, semantic integration, and self-monitoring processes ^66–69^. Notably, cortical thickness yielded no significant regional associations with symptoms, and disorganization was the one dimension that did not contribute significantly. This dissociation suggests that symptoms are more closely linked to how cortical thickness is organized across the brain regions than to where it is reduced. In other words, the relational structure of thickness covariance, captured by gradient position and small-world topology, carries symptom-relevant information that is not apparent in univariate thickness differences between patients and controls. This finding aligns with the view that psychopathology reflects distributed disruptions in cortical network architecture rather than focal morphological differences ^25,70,71^, and underscores the added value of network-level analysis of structural MRI beyond conventional alteration mapping.

Finally, using a network-based clustering framework, we identified two robust schizophrenia subtypes characterised by divergent anterior cingulate-posterior profiles of cortical thickness alteration, embedded within the macroscale gradient-topology organisation of the cortex. Critically, although S1 and S2 differed in global mean cortical thickness, the spatial pattern distinguishing the two subtypes was virtually identical after controlling for mean thickness (*r* = 0.997), demonstrating that the subtype difference reflects a genuine divergence in the topographic distribution of thinning rather than an overall severity gradient. Neuroanatomically, S2 was characterised by widespread cortical thinning spanning frontal, temporal, parietal, and central cortices with relative preservation of the ACC, whereas S1 showed a more diffuse whole-cortex thinning pattern with pronounced ACC involvement. Clinically, S1 showed an age-related trajectory of increasing positive and disorganization symptoms, potentially reflecting the persistent ACC network disruption observed in this subtype, whereas S2 did not show this age-related increase.

Data-driven subtyping of schizophrenia has been pursued across multiple methodological approaches ^12,17,72,73^, including machine learning applied to regional morphometry and sequential staging models such as SuStain, which have identified frontal- and temporal-hippocampal-anchored patterns of cortical alteration ^12,17,73^. Our findings extend this literature by showing that subtype heterogeneity is organised along the low-dimensional eigenmodes of cortical organization, with the two subtypes reflecting differential weighting of disruptions along the principal gradient-topology axis rather than spatially arbitrary regional abnormalities. This distinction has methodological implications: staging-based approaches such as SuStain ^12,74^ infer subtypes by ordering abnormalities into hypothetical disease trajectories, assuming monotonic progression, an assumption that remains contested in schizophrenia given its non-linear and heterogeneous illness course ^2,4^. Our gradient-topology framework identifies subtypes based on spatial structure alone, making it robust to the absence of monotonic staging. Future longitudinal studies could directly test whether disease processes unfold along the same network organisational axes identified here, and whether this predicts convergence with staging-based subtyping approaches. Given that cortical gradients capture neurodevelopmental differentiation into specialised systems ^75,76^, our findings suggest that schizophrenia subtypes may arise from complementary deviations along this organisational axis, ultimately converging toward a shared system-wide alteration pattern. Together, these results advance a network-level reinterpretation of schizophrenia heterogeneity beyond region-centric frameworks.

### Limitations and future directions

While this large-scale international multisite data provide robust and replicable evidence for systematic disruption of cortical network architecture in schizophrenia, the cross-sectional design limits direct mechanistic inference. In particular, population-based developmental cohorts, combined with detailed genetic and environmental risk phenotyping, are critical for elucidating how deviations in cortical network organization emerge prior to illness onset. Such approaches are well suited to disentangle neurodevelopmental processes shaping large-scale cortical architecture from later disease-related network alterations. Complementary, future work in clinical high-risk and early psychosis cohorts may advance how the observed disruptions in gradient and small-world topology manifest closer to the disease phenotype, enabling investigation of early pathological mechanisms within the illness trajectory. In parallel, large-scale genetic resources, including polygenic risk scores, can be leveraged to characterize the genetic architecture of cortical network organization and its overlap with schizophrenia risk. Together, these complementary longitudinal strategies will help bridge normative development, genetic liability, and disease-related altered cortical network organization, advancing mechanistic understanding of schizophrenia at the network level.

### Conclusions

In summary, this study leveraged large-scale, multisite neuroimaging data from the ENIGMA Schizophrenia Working Group to provide robust evidence for systematic disruption of cortical gradient and network topology in schizophrenia. At the level of macroscale organization, schizophrenia was characterized by widespread alterations in cortical thickness covariance gradients, reflecting system-level reorganization of cortical architecture. In parallel, disruptions in small-world network topology were more spatially localized. Critically, both gradients and topology revealed a gradedbrain-symptom association, in which symptom dimensions contributed along a spectrum from disorganization and hostility through negative symptoms to affective symptoms, a pattern that was specific to cortical network organization and distinct from the cortical thickness association pattern. This suggests that cortical network measures capture symptom-relevant information that conventional thickness mapping does not. By integrating gradients and topology, we further identified two robust cortical subtypes with stable anterior-posterior differentiation, supporting the notion that schizophrenia heterogeneity is embedded within large-scale network organization and potentially shaped by developmental and genetic factors. Notably, the subtype with anterior cingulate involvement showed greater positive symptom burden at older ages despite no greater overall cortical thinning, a dissociation that warrants longitudinal confirmation. These findings highlight the complementary roles of cortical gradients and network topology in capturing both shared organizational alterations and individual symptom variability, offering a framework for how macroscale cortical organization shapes clinical heterogeneity in schizophrenia.

## Data availability

The neuroimaging data used in this study were obtained through the ENIGMA consortium and are available in accordance with ENIGMA data-sharing policies. **Supplementary Materials** for the figures and region-specific statistics are provided with this paper.

## Code availability

All the analytic scripts and visualization are openly available at a GitHub repository (https://github.com/wanb-psych/enigma_scz). The python packages are completely open for use, see documentations: BrainSpace (https://brainspace.readthedocs.io/en/latest/) for gradients construction, NetworkX (https://networkx.org/) for small-world topology construction. Scikit-learn (https://scikit-learn.org/stable/) for PLS construction, Statsmodels (https://www.statsmodels.org/stable/index.html) for the statistics, and ENIGMA toolbox (https://enigma-toolbox.readthedocs.io/en/latest/) for spatial autocorrelation null models (spin test).

## Acknowledgements

The HUBIN cohort was supported by the Swedish Research Council (grant numbers: K2004-21X-15078-01A, 2006-2992, 2006-986, and 2008-2167, K2010-62X-15078-07-2, K2012-61X-15078-09-3), the Söderström-Königska Foundation, the Knut and Alice Wallenberg Foundation, the regional agreement on medical training and clinical research between the Stockholm Region and the Karolinska Institutet, and the HUBIN project. The NU cohort was supported by NIH grants P50 MH071616, R01 MH056584, R01 MH084803 (Wang PI), T32 NS047987 (Cobia PI), U01 MH097435 (Wang, Turner, Ambite, Potkin PIs), R01 EB020062 (Miller, Paulsen, Mostfosky, Wang PIs), NSF 1636893 (Pestilli, Wang, Saykin, Sporns PIs), NSF 1734853 (Pestilli, Garyfallidis, Henschel, Wang, Dinov PIs). The Voices cohort was supported by an NHMRC project grant and the authors acknowledge the facilities and scientific assistance of the National Imaging Facility, a National Collaborative Research Infrastructure Strategy (NCRIS) capability, at the Swinburne Neuroimaging Facility, Swinburne University of Technology. The Galway cohort was supported by grants from the European Research Council (grant no. 677467), Science Foundation Ireland (16/ERCS/3787), and the Health Research Board (RL - 2020 -007) to Gary Donohoe. The COBRE cohort was supported by a NIH COBRE Phase I grant (1P20RR021938, Lauriello, PI and 2P20GM103472, Calhoun, PI) awarded to the Mind Research Network. The ESO cohort was supported by the Ministry of Health of the Czech Republic, grant NU22-04-00143 and grant from the Program Johannes Amos Comenius under the Ministry of Education, Youth and Sports of the Czech Republic CZ.02.01.01/00/23_020/0008560. The IGP cohort was funded by Project Grants from the NHMRC (#630471 and #1081603) and the Macquarie University’s ARC Centre of Excellence in Cognition and its Disorders (CE110001021). The MCIC cohort was supported by the National Institutes of Health (NIH/NCRR P41RR14075 and R01EB005846 (to Vince D. Calhoun)), the Department of Energy (DE-FG02-99ER62764), the Mind Research Network, the Morphometry BIRN (1U24, RR021382A), the Function BIRN (U24RR021992-01, NIH.NCRR MO1 RR025758-01, NIMH 1RC1MH089257 to Vince D. Calhoun), the Deutsche Forschungsgemeinschaft (research fellowship to Stefan Ehrlich), and a NARSAD Young Investigator Award (to Stefan Ehrlich). The KaSP cohort was supportred by the Swedish Medical Research Council (SE: 2024-02812, 2023-02827, 2017-00875, 523-2014-3467); the Swedish Brain Foundation (SE: FO2023-0333); Karolinska Institutet and Stockholm County Council (20160328, 20180487, 20190175, 20190447); Torsten Soderberg Stiftelse; The Swedish Medical Society, and Erling Persson Foundation. Tilo Kircher receives funding from the German Research Foundation (DFG) FOR 2107, SFB/TRR 393 (“Trajectories of Affective Disorders”, project grant no 521379614), and the Germany’s Excellence Strategy (EXC 3066/1 “The Adaptive Mind”, Project No. 533717223), as well as the DYNAMIC center, funded by the LOEWE program of the Hessian Ministry of Science and Arts (grant number: LOEWE1/16/519/03/09.001(0009)/98). Biosamples and corresponding data were sampled, processed and stored in the Marburg Biobank CBBMR. Udo Dannlowski was funded by the German Research Foundation (DFG, grant FOR2107 DA1151/5-1, DA1151/5-2, DA1151/9-1, DA1151/10-1, DA1151/11-1 to UD; SFB/TRR 393, project grant no 521379614) and the Interdisciplinary Center for Clinical Research (IZKF) of the medical faculty of Münster (grant Dan3/016/26). Matthias Kirschner acknowledges funding from the Swiss National Science Foundation (P2SKP3_178175 and grant number 32003B_219240).

## Authors’ contributions

**Conceptualization:** B.W., M.K. **Methodology:** B.W., M.K. **Formal analysis:** B.W. **Writing - Original Draft:** B.W., M.K. **Writing - Review & Editing:** B.W. V.W., R.A.I.B, S.L., C.A.M, Y.S.F, Y.H., ENIGMA Schizophrenia Working Group, S.K., P.M.T, T.G.M.E, J.A.T, B.C.B, S.L.V, M.K. **Visualization:**B.W. **Data curation:** B.W., M.K., ENIGMA Schizophrenia Working Group. **Project administration:** B.W., M.K. **Funding acquisition:** M.K. **Supervision:** B.W., M.K.

