## Supplementary Fig. for "Individualized cortical gradient and network topology reveal symptom-linked disruptions and neurobiological subtypes in schizophrenia"

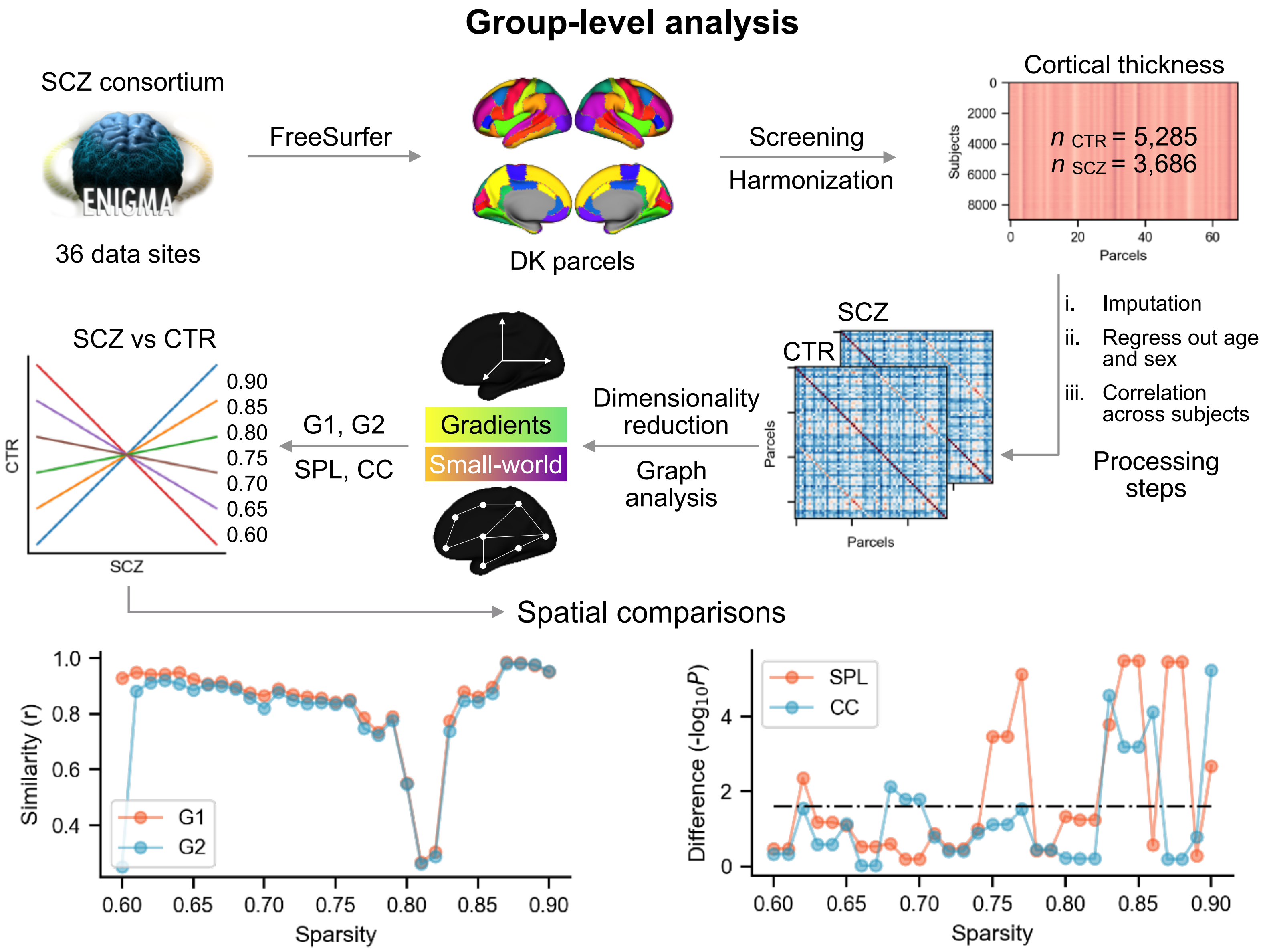


**Fig. S1: Gradients and small-world topology calculated from group-level covariance analysis across subjects in schizophrenia and controls.**

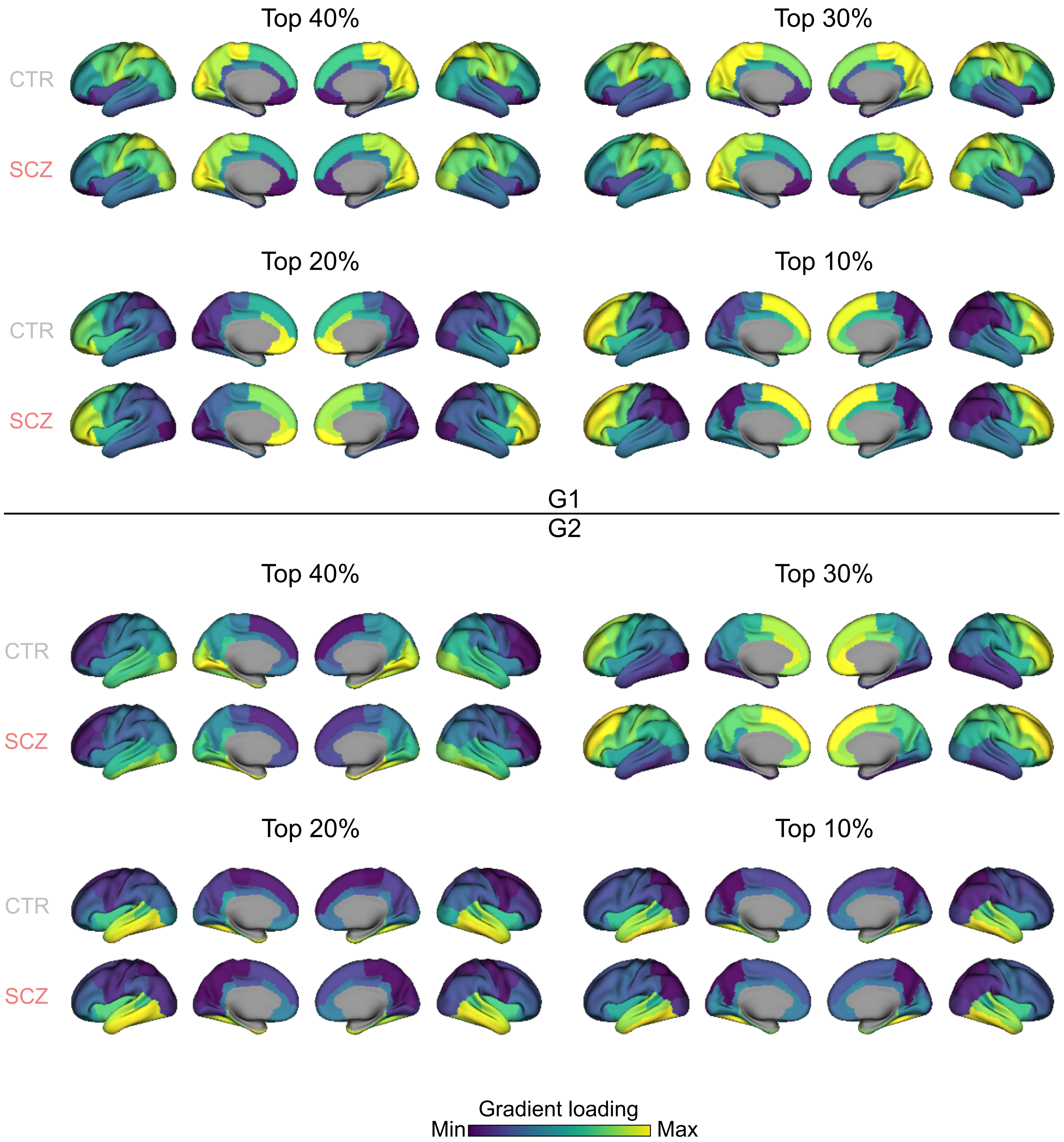


**Fig. S2: Group-level small-world topology maps using top 10%, 20%, 30%, and 40% threshold for the covariance matrix.**

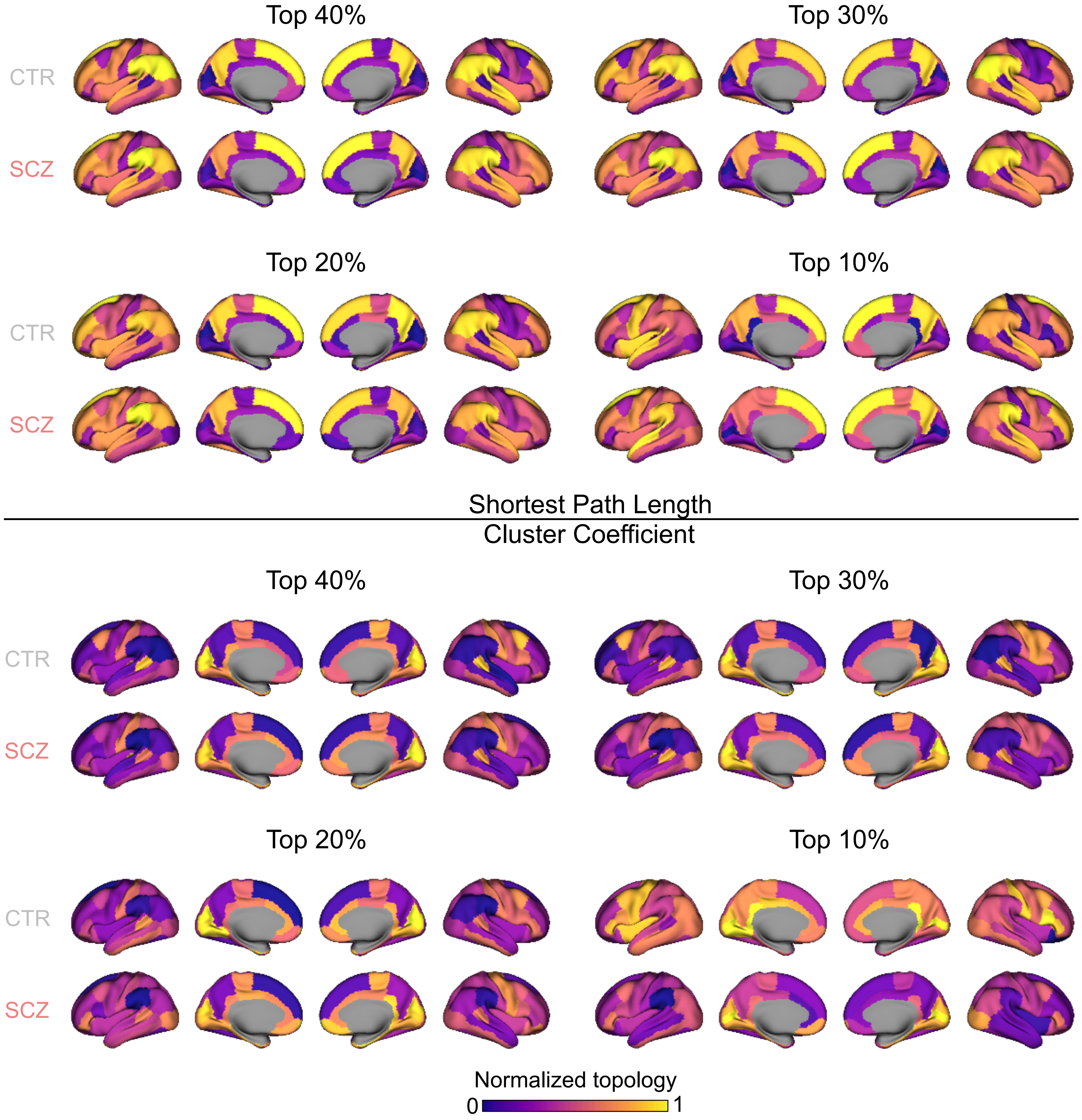


**Fig. S3: Group-level small-world topology maps using top 10%, 20%, 30%, and 40% threshold for the covariance matrix.**

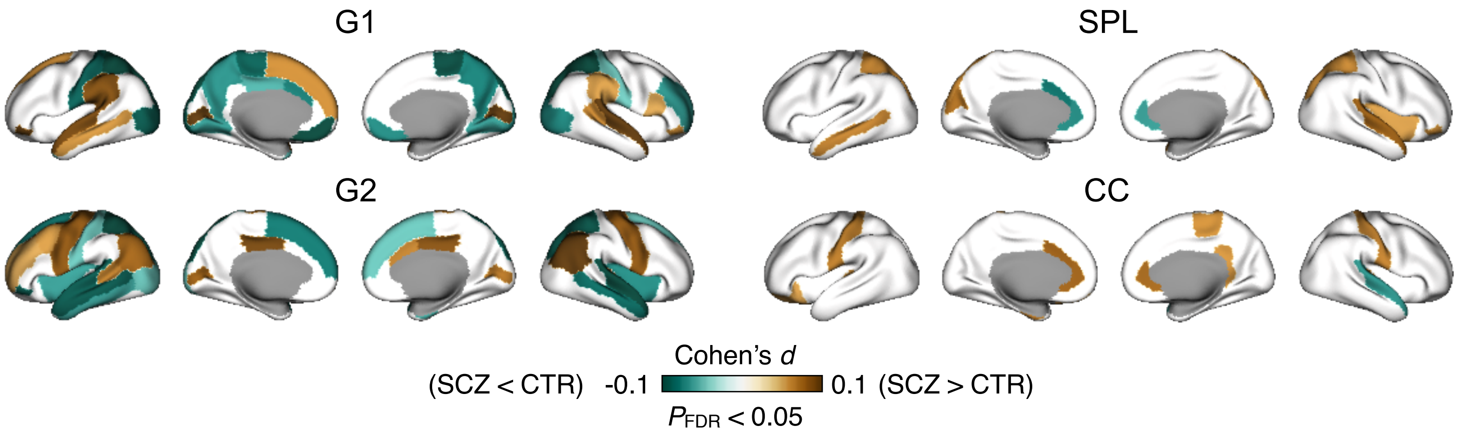


**Fig. S4: Robustness analysis for group comparisons in Fig. 2B with adding into intracranial volume (ICV) into the regression model.**

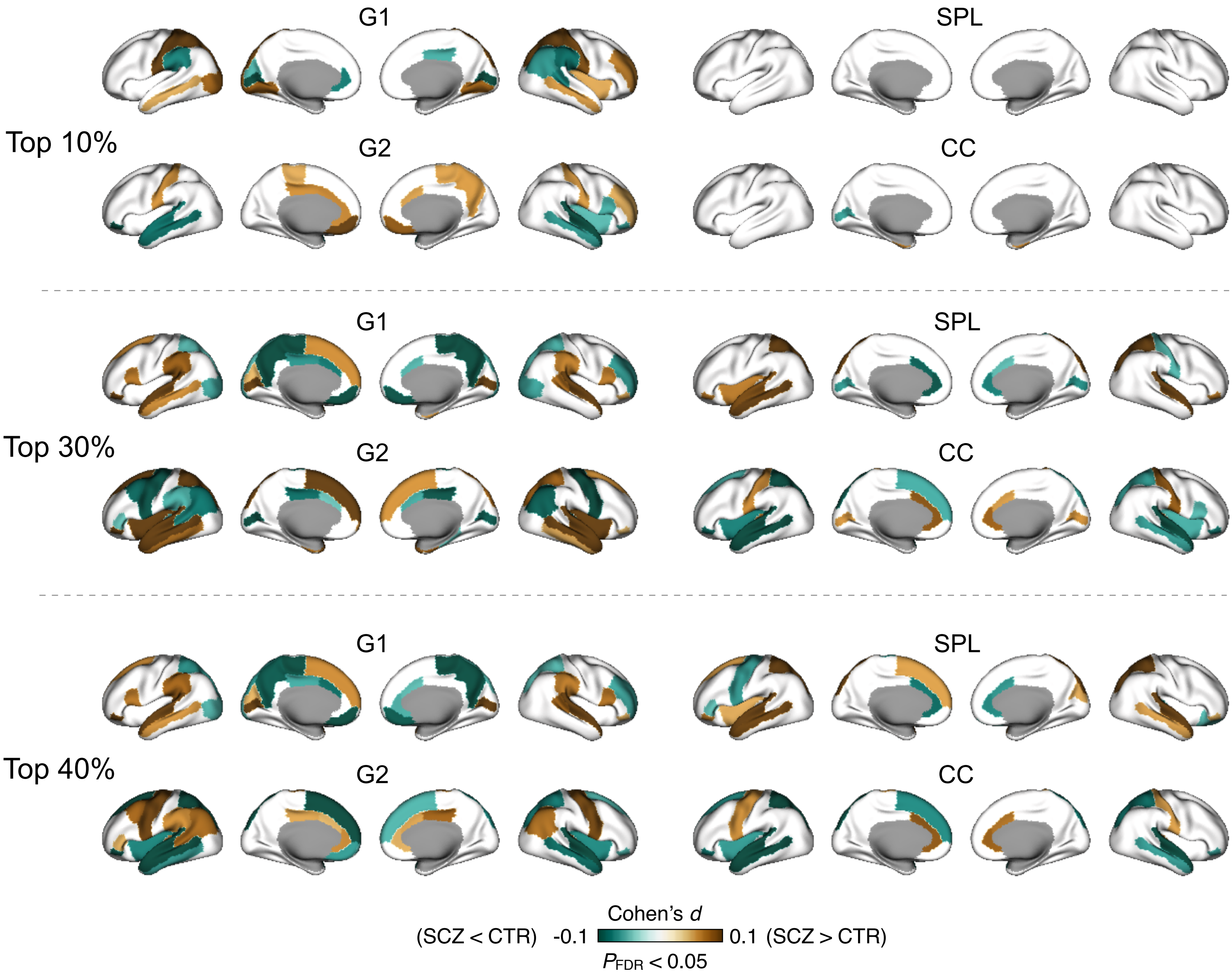


**Fig. S5: Robustness analysis for group comparisons in Fig. 2B using different top covariation thresholds of 10%, 30%, and 40%.** The main results in Fig. 2B used top 20%.

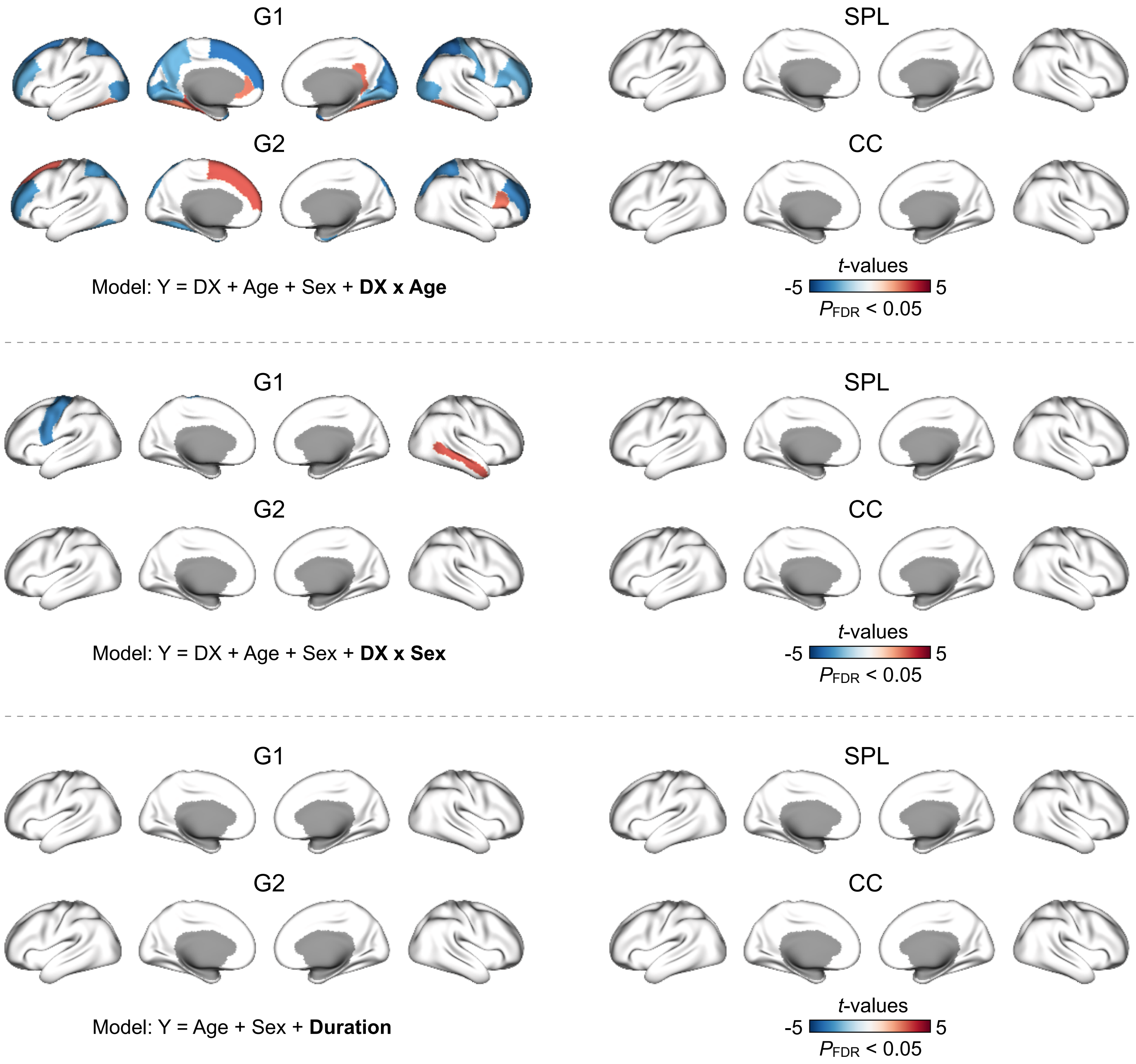


**Fig. S6: Interaction and disease supplementary analysis for Fig. 2B.** Maps of interaction between age and diagnosis (DX), interaction between sex and DX, and effects of disease duration were shown in this figure.

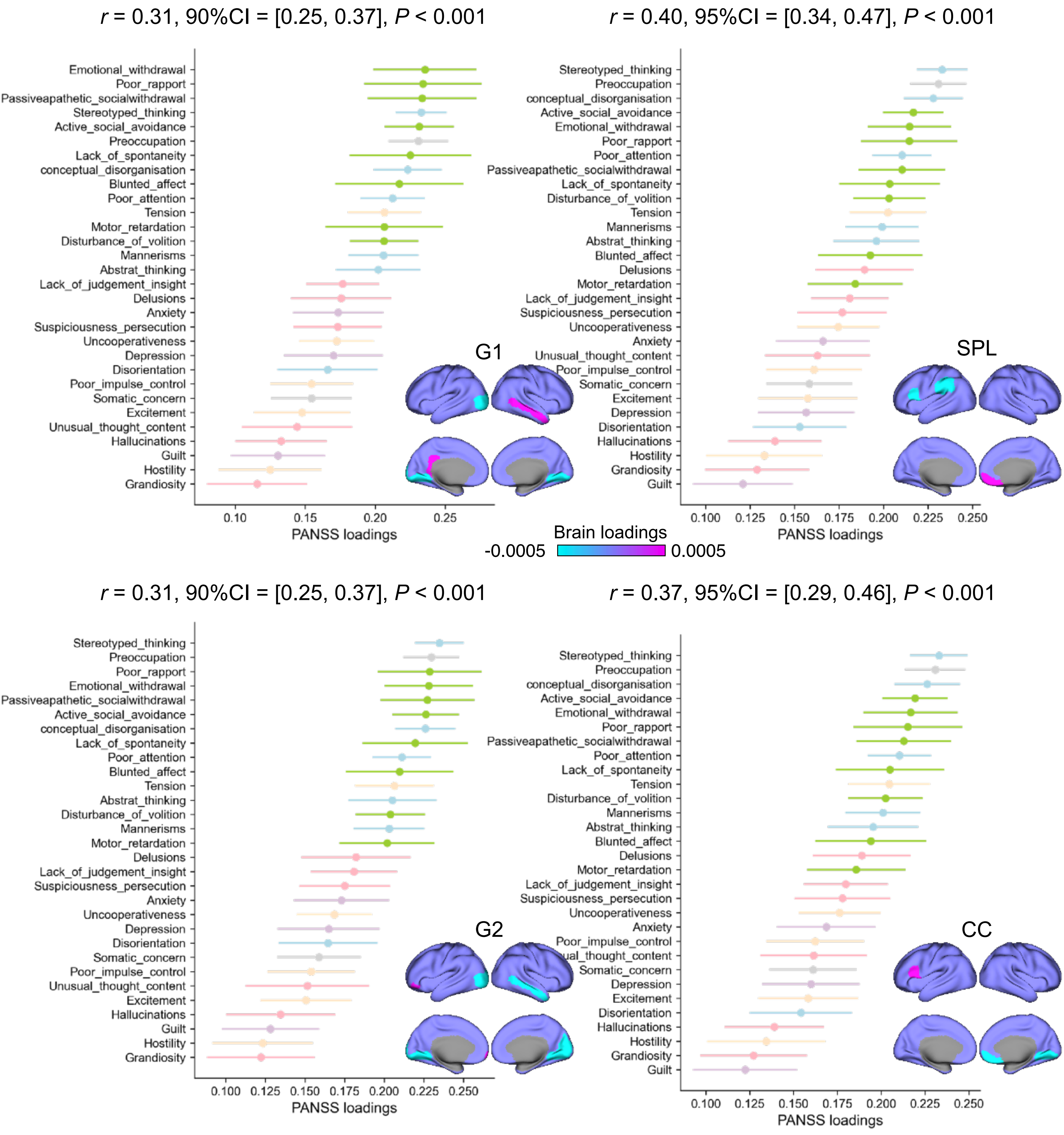


**Fig. S7: Item-based PLS analysis between clinical symptoms and brain features.**

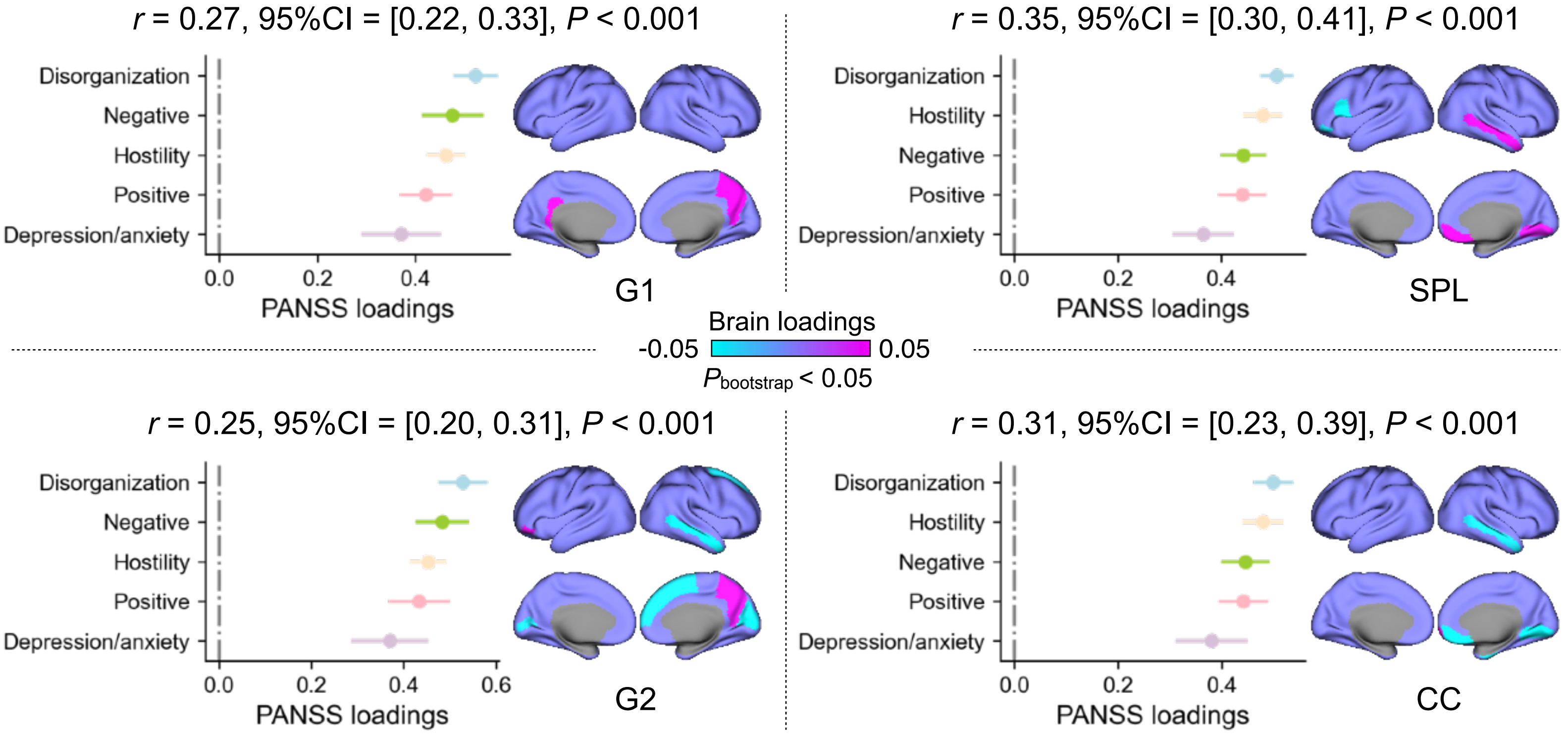


**Fig. S8: Factor-based PLS analysis between clinical symptoms and brain features with regressing age and sex, compared to Fig. 3 (regressing age, sex, and disease duration).**

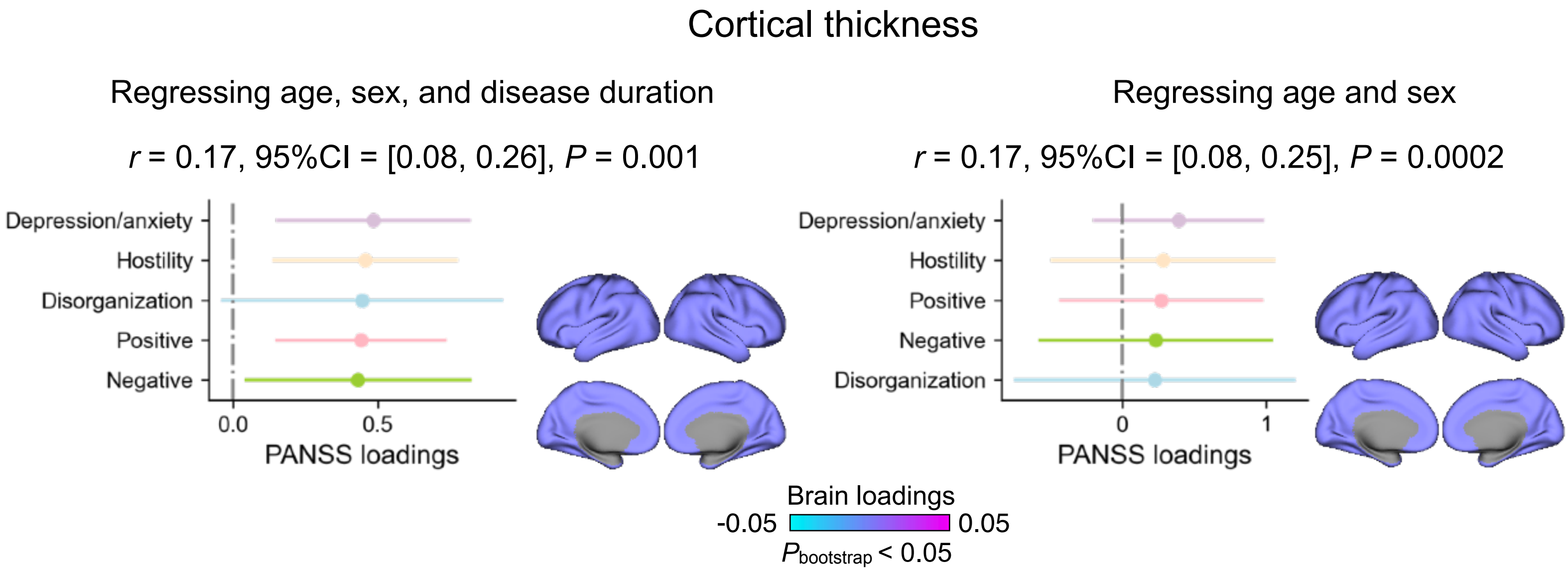


**Fig. S9: Factor-based PLS analysis between clinical symptoms and cortical thickness.**

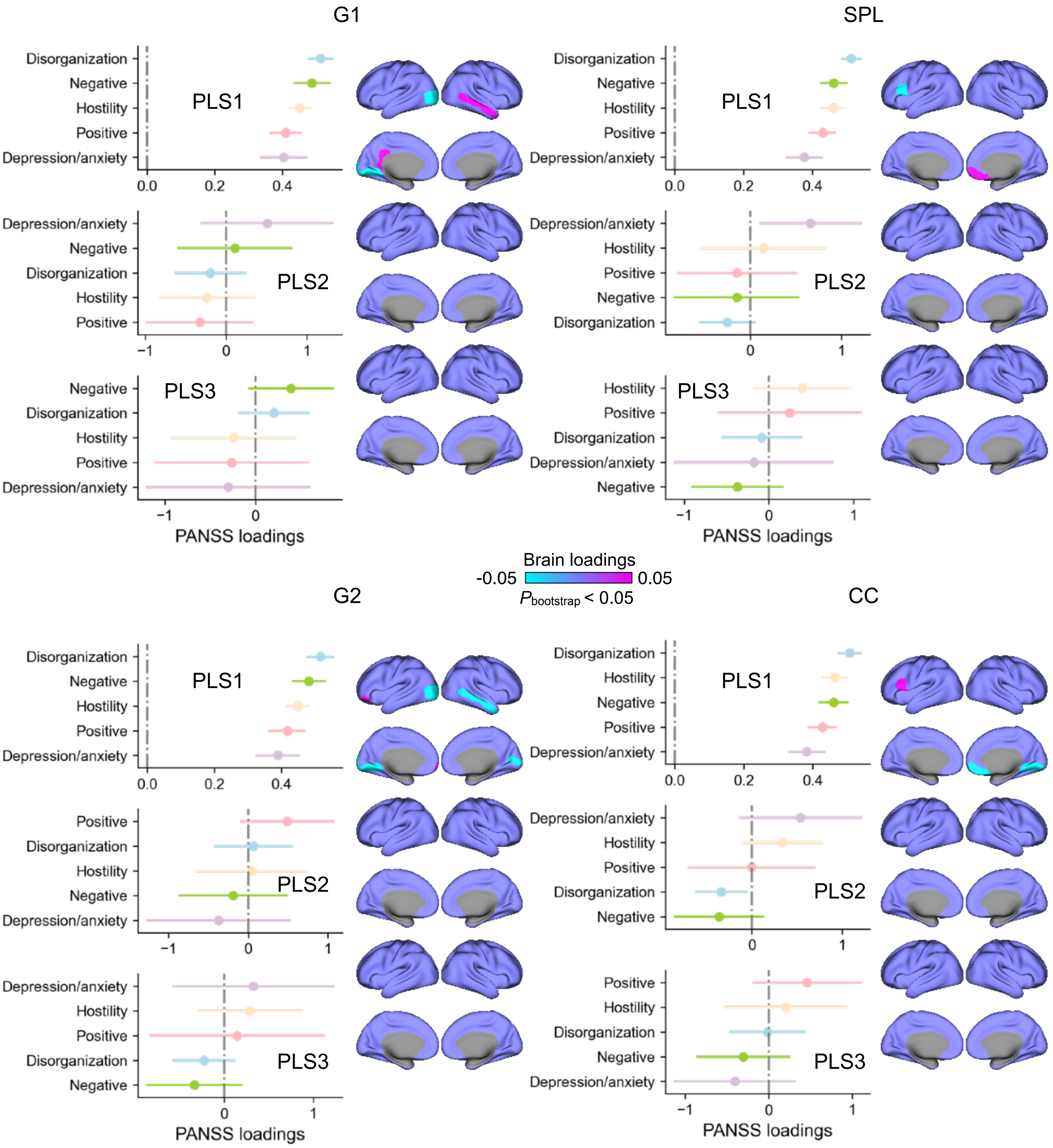


**Fig. S10: Factor-based PLS analysis (3 components) between clinical symptoms and brain features with regressing age, sex, and disease duration, compared to Fig. 3 (1 component).**

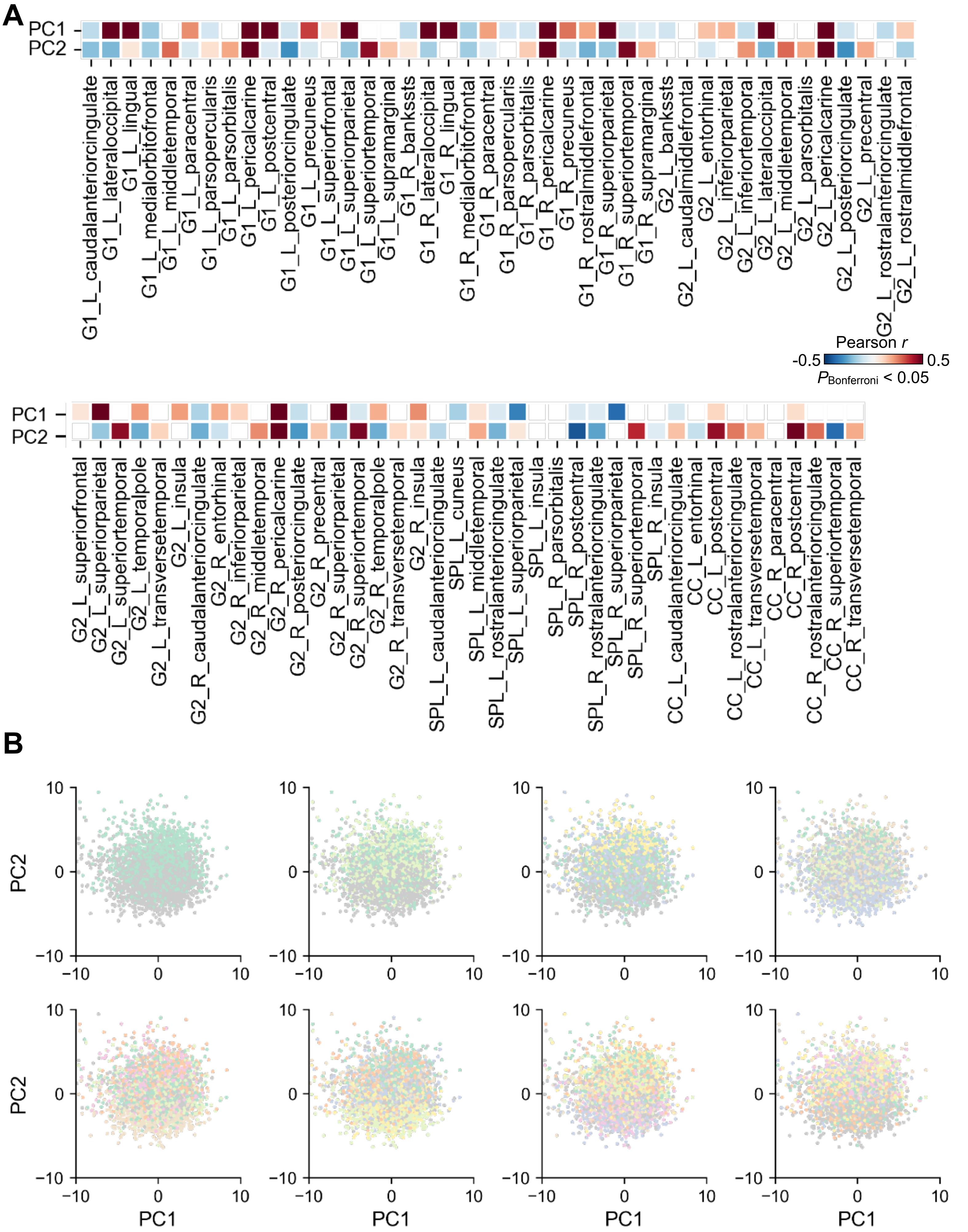


**Fig. S11: Supplementary figure for the PCA and subtpying in Fig. 3. A**: Correlation between 79 brain features and the first two PCs. **B**: Subtyping solutions from 2 to 9 along the first two PCs.

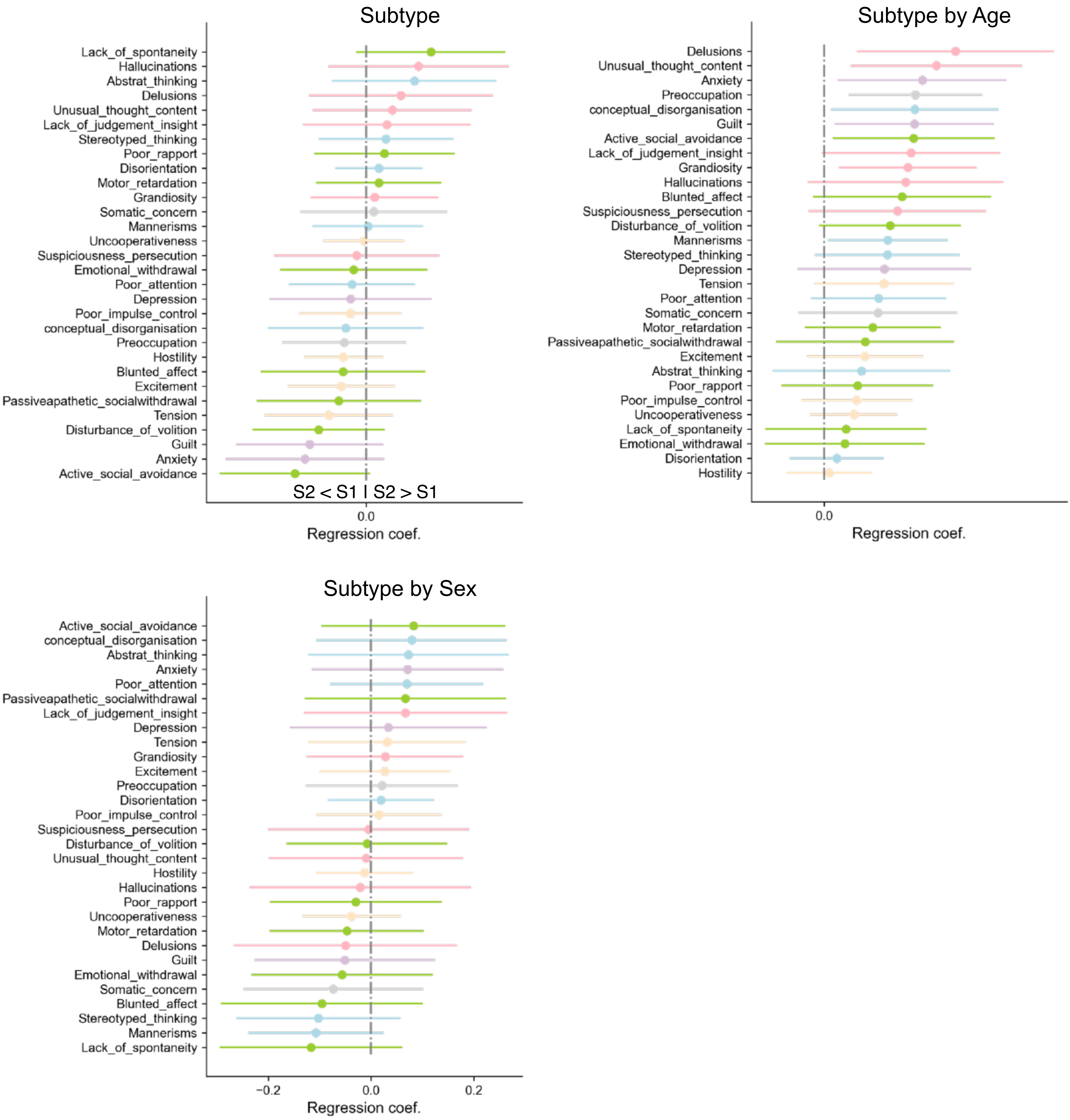


**Fig. S12: Clinical item-based difference between subtype 1 and subtype 2.**

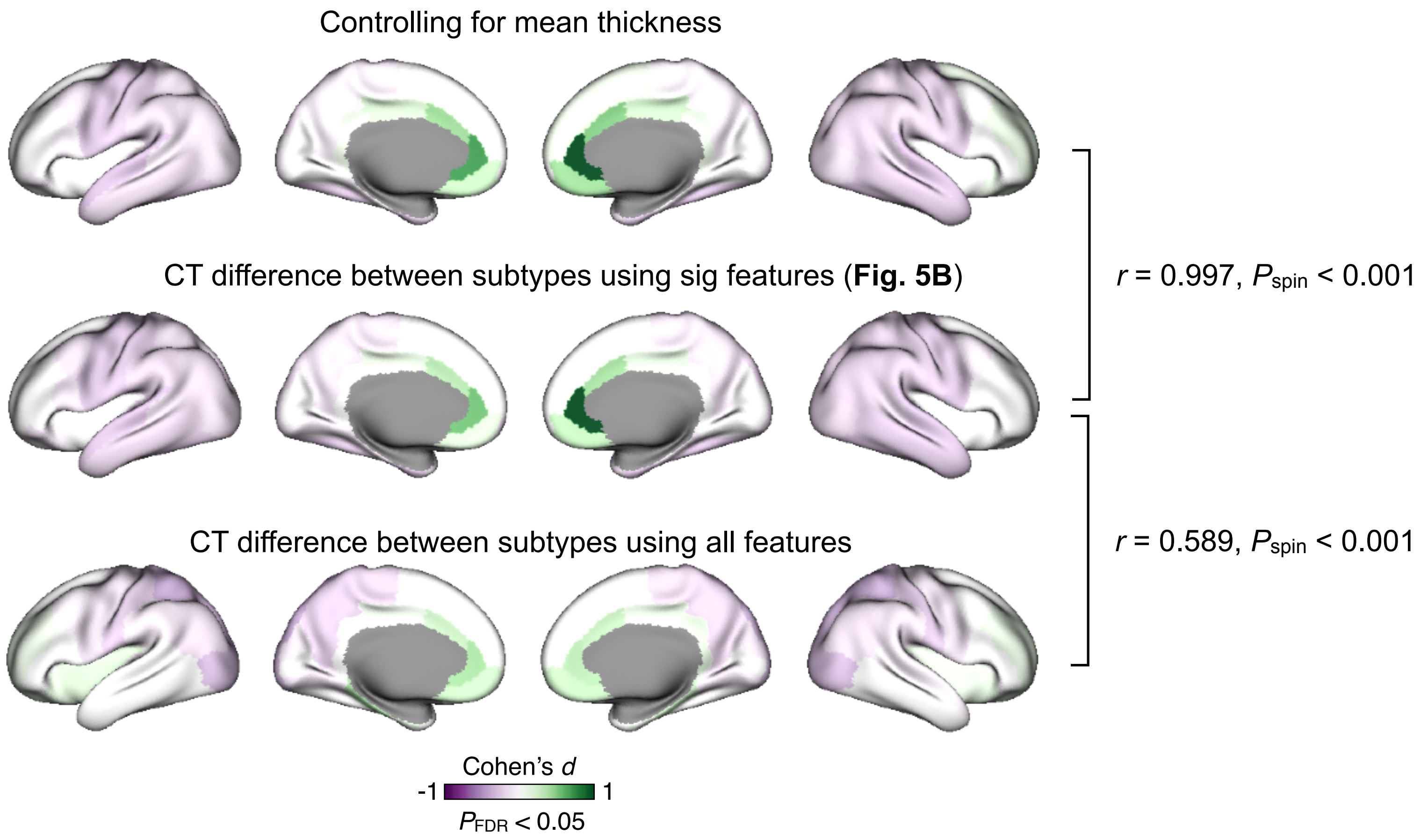


**Fig. S13: Robustness test of cortical thickness difference between subtype 1 and subtype 2.**

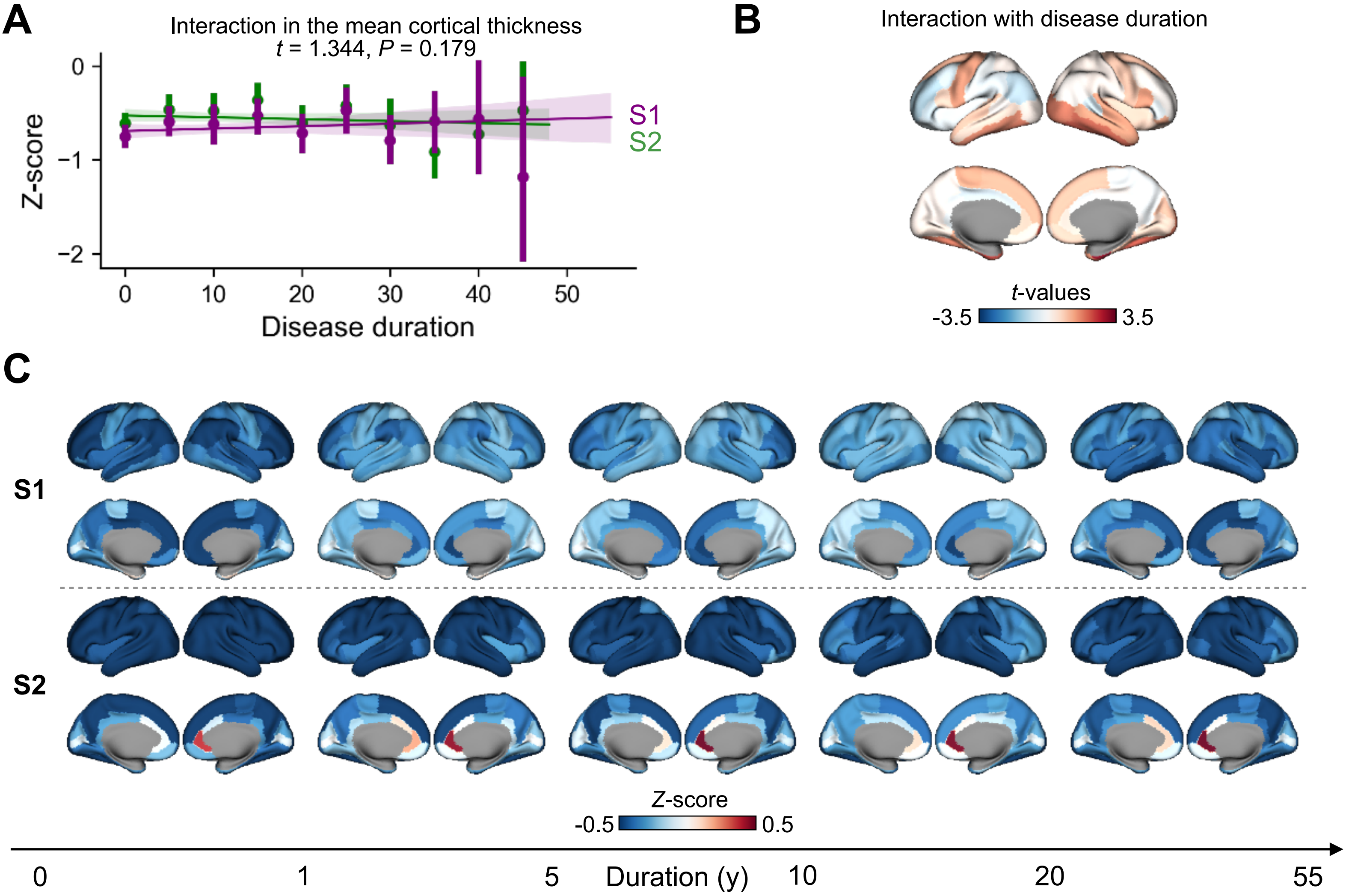


**Fig. S14: Z-score maps in S1 and S2 in 5 disease duration groups from 0 to 55. A:** Interaction between subtype and disease duration in the mean cortical thickness. We did not observe significant interaction. **B:** Interaction between subtype and disease duration in the region level cortical thickness. It showed no regions surviving from statistical multiple comparisons correction. **C:** Regional mean z-score maps in S1 and S2 along disease duration.

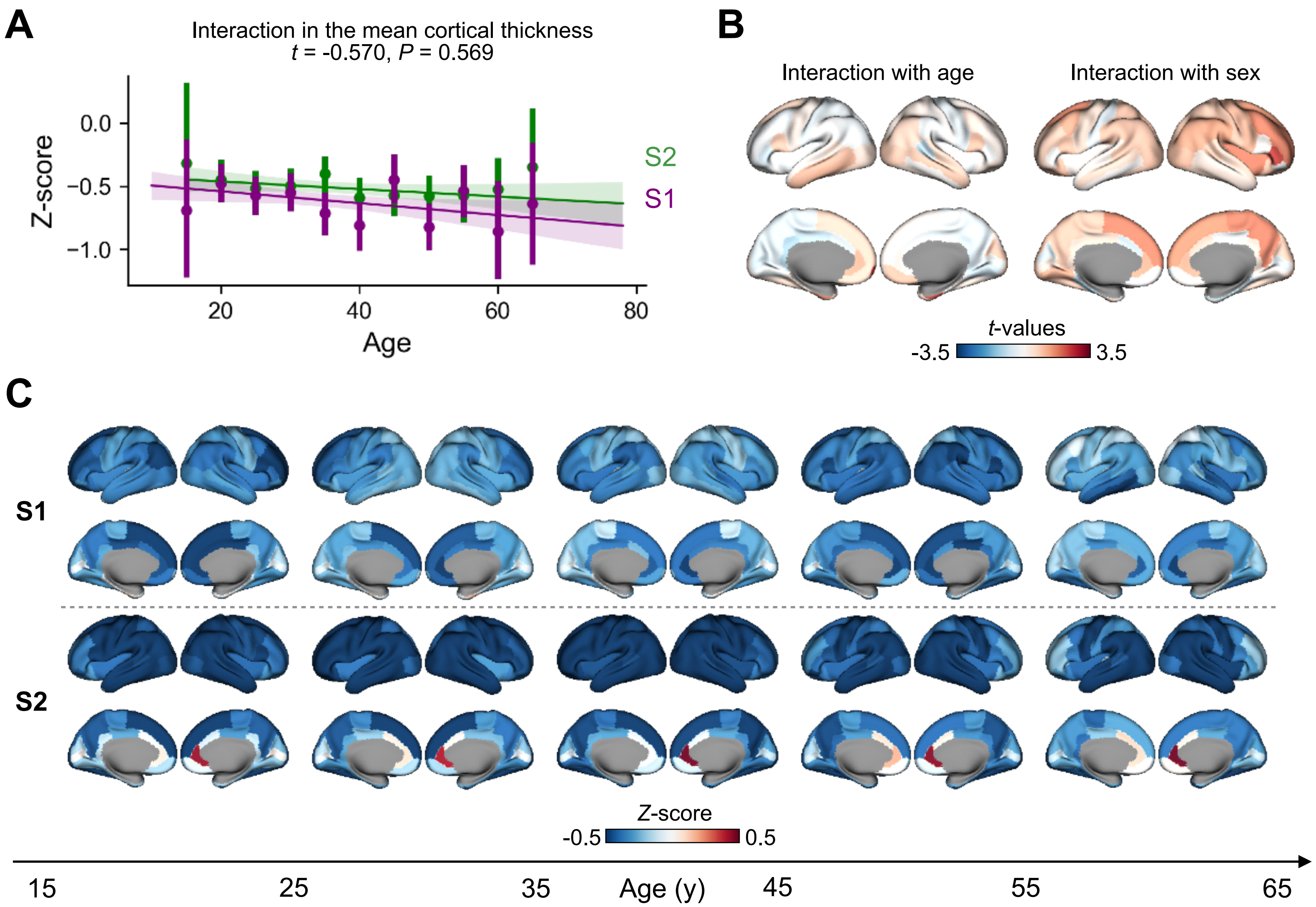


**Fig. S15: Z-score maps in S1 and S2 in 5 age groups from 15 to 65. A:** Interaction between subtype and age in the mean cortical thickness. We did not observe significant interaction. **B:** Interaction between subtype and age/sex in the region level cortical thickness. It showed no regions surviving from statistical multiple comparisons correction. **C:** Regional mean z-score maps in S1 and S2 along age.
